# Integrative proteomics and network biology approach to identify potential urine based biomarkers for tuberculosis

**DOI:** 10.1101/2023.05.19.23289652

**Authors:** Sangeetha Subramaniam, Ankur Varshney, Rupak Singla, Digamber Behera, Ranjan Kumar Nanda

## Abstract

Urine based biomarker discovery employing proteomics platform has been successfully attempted for multiple diseases. Urine is an excellent source of biomarker discovery but its potential is not fully tapped in tuberculosis (TB) diagnostics. In the present study, proteomic profiling of urine samples from thirty five subjects (Mean age=41 years (15-76), M/F=28/7) belonging to active TB, latent TB, lung cancer, chronic obstructive pulmonary disorders (COPD) and healthy subjects were carried out employing a robust multiplex technique. We identified 131 proteins out of which 16 molecules showed at least two-fold change in TB. The study identified a signature of three putative markers, leucine-rich alpha-2-glycoprotein (up-regulated), roundabout homolog 4 and isoform 2 of prostatic acid phosphatase (down-regulated) that could differentiate active TB from other pulmonary diseases. Besides, we investigated whether a network based approach can be efficiently used to expand dynamic coverage, gain a comprehensive view of underlying perturbed functions during the infection and to discover potential biomarkers. While comparing the functionally associated sub-networks of active TB with healthy urine proteome, we identified 54 proteins from the discriminative TB sub-network, some of which are known to be involved in the infection process. Few examples in this study like serpin peptidase inhibitor and catenin that has not been identified in the experiment but detected in the difference network demonstrate that proteomic profiling when integrated with network biology method could be a holistic approach to expand the dynamic range and identify potential candidate biomarkers and also provide a broad overview of perturbed functions during the infection.

## Introduction

According to the 2022 WHO report 21.4 lakh fresh cases of tuberculosis (TB) has been reported in India (1). The global menace continues to prevail despite the arsenal of drugs, but again these are mostly effective only when detected early and treated adequately. Early diagnosis of TB by identification of protein biomarkers is quite challenging, but holds enormous potential to prevent complications and save a subpopulation with the effective disease management. Pathological events mediated by structural and functional biochemical changes in the metabolic system get reflected by the differential activity of specific marker molecules. Biomarker discovery capitalizes on this principle of comparing the physiological states, phenotypes and their differential activity between control and disease states. The application of biomarker for diagnostic tool development has been widely investigated in several diseases including TB (2), but understandably requires rigorous statistical consideration and validation pipeline, to ensure high sensitivity and specificity.

Non-invasive nature, ease of collection, availability in large volume, convenience of repeated sampling, and stability features of urine makes it an attractive matrix for biomarker discovery. Urine based biomarkers have been reported for several disease conditions (3). The diagnostic potential of urine for TB has constantly been explored using different approaches (4, 5). Earlier reports showed that lipoarabinomannan (LAM) urine assay has suboptimal sensitivity (6) for routine clinical use and their evaluation for paediatric TB and advanced HIV TB diagnosis are yet to be confirmed (7). Several standard methods are recommended to optimise the urine based clinical diagnosis (8, 9); however, the loss of potential marker molecules that lie below limit of detection (LOD) and the low dynamic range are still inevitable. The latter remains the inherent shortcoming of proteomics and renders the biomarker identification analogous to search of needles in haystacks especially amongst the high abundance proteins like albumin and uromodulin. The molecules, that often elude the experiment possibly due to their low abundance and their masking by high abundant molecules have been the key players of pathological events and detected as potential markers in some cases (10).

Network analysis have been applied to several biological problems and has aided the investigation of several disease conditions (11). We investigate here the application of a simple network based method to expand the dynamic range of proteomic studies and also to identify putative biomarkers. First, we profile the urine proteome from active TB patients using isobaric tag for relative and absolute quantification (iTRAQ), and compare it with the a *priori* healthy urine proteome dataset by constructing their functionally associated protein interaction network. The discriminative sub-network enumerates a set of putative marker molecules that presumably eluded the experiment and involved in assisting or resisting *Mycobacterium tuberculosis* (Mtb) and hence provide new insights about the pathogenesis. We also investigated the proteomic content of urine samples from other disease conditions like lung cancer, chronic obstructive pulmonary disease (COPD) and latent TB to decipher specificity of these markers by their differential expression. In this study we report a set of putative markers for TB that are at exploratory stage and needs further validation using an independent set of patient samples.

## Experimental procedures

### Patient Enrolment

All study participants recruited were TB suspects attending the outpatient department at Lala Ram Swarup Institute of Tuberculosis and Respiratory Diseases (LRSI) in New Delhi. Each participant underwent clinical examination and laboratory assessment for TB. Recruited subjects were assessed for medical history, cigarette smoking, body-mass index, tuberculin skin test and medication status. Demographic details with clinical characteristics are presented in Table 1. Healthy relatives of TB patients and donors from International Centre for Genetic Engineering and Biotechnology (ICGEB), New Delhi were enrolled as controls. Healthy controls were asymptomatic individuals and underwent tuberculin skin test (Mantoux test) and full diagnostic assessment to exclude TB. Due to logistic challenges, tuberculin skin test results were considered for defining latent TB infection (LTBI). A person having more than 10 × 10 mm skin indurations and otherwise showing no evidence of active TB was defined as LTBI. Sputum microscopy +ve subjects were included as active TB patients. To allow systematic inflammatory processes as reference cohorts that can mimic TB, we recruited patients with other symptomatic overlapping clinical presentations like lung cancer and COPD by experts from the collaborative hospital. Exclusion criteria for all study groups were inability or unwillingness to give written informed consent by the patients or parents, age under 14 years, co-infection with HIV and compromised immune status. Fully informed written consent was obtained from every recruit in accordance with local and institutional ethics committees’ policy. Study protocol and consent forms were approved by the institutional review boards of ICGEB and LRSI. Patients with suspected TB who were eligible and declined not to participate were not enrolled for the study.

**Table 1:** Epidemiological details of recruited subjects (n=35) used in Urine proteomics biomarker study. M: Male, F: Female, no Info: no information, TB: Tuberculosis, PPD status is based on minimum 10×10 mm, COPD: Chronic obstructive pulmonary disorder

|  | Discovery set (iTRAQ-1) |  |  |  |  |
| --- | --- | --- | --- | --- | --- |
|  | TB | PPD +ve | PPD -ve | COPD | Lung Cancer |
| **Subject numbers** | 10 | 10 | 5 | 5 | 5 |
| **Mean age (Range) in years** | 33(20-55) | 32(23- 60) | 31(15- 45) | 60(52- 65) | 64(45-76) |
| **Gender (M/F)** | (7/3) | (8/2) | (4/1) | (5/-) | (4/1) |
| **PPD (+ve/-ve/no info.)** | 9/1/- | 10/-/- | -/5/- | -/-/10 | -/-/10 |
| **Cough (yes/no)** | 10/- | 3/7 | 1/4 | 5/- | 4/1 |
| **Expectoration (yes/no)** | 8/2 | -/10 | -/5 | -/5 | -/5 |
| **Chest pain (yes/no)** | 6/4 | 1/9 | 1/4 | 2/3 | 4/1 |
| **Abnormal X-ray (yes/no/no info.)** | 8/2/- | -/10/- | -/5/- | -/-/10 | -/-/10 |
| **Cavity (yes/no/no info.)** | 3/7/- | -/10/- | -/5/- | -/-/10 | -/-/10 |
| **Smear (+ve/-ve/no info.)** | 10/-/- | -/10/- | -/5/- | -/-/10 | -/-/10 |

### Urine Sample Collection and Processing

Mid-stream, spot urine samples were collected using chilled sterile falcon tubes of 50 mL capacity. After collection, urine samples were packed in three layered plastic containers with adsorbent to transport it to research institute at 4 °C within 2-3 hours of sample collection. Protease inhibitors (per 50ml); 100mM of sodium azide, 100 µL of phenyl methyl sulfonyl fluoride (PMSF; 2%) and 1 µL of leupeptin (100mM) were added to the urine samples. Sample was aliquoted in 15 ml falcon tubes to store at −80 °C and the whole processing was completed within 4-5 hours of sample collection from recruited subjects. These samples were treated with no more than two freeze thaw cycles before undertaking further processing for proteomics study. Before undertaking the experiment, urine samples from the internal institutional biobank were thawed in ice to reach 4 °C and then centrifuged at 10,000 × g for 20 mins at 4 °C to remove the cells, precipitated materials and debris. Subsequently clear supernatant extracted from urine was lyophilised and dissolved in minimum amount of phosphate buffer saline (100 mM, pH 7.4). Equal amount of proteins were pooled from five subjects with similar age and gender attributes from each diseases groups to undertake proteomics study. A part of these samples have been used in our previous study where we investigated the volatile organic compounds (VOCs) to aid TB biomarker discovery (4).

### Quantitative Proteomics

Multiplexing technique was adopted to relatively quantify urine proteins from five separate study groups in single iTRAQ experiment (Figure 1). Biological replicates for active and latent TB groups were included in this study. Tryptic peptides were generated from 100 μg of proteins by trypsinization of the samples, reduced and MMTS cys alkylated peptides and further labelling were conducted using isobaric tags for relative and absolute quantification (iTRAQ) chemistry following protocol provided by the manufacturers (Applied Biosystems, USA). The isobaric tagged peptides were pooled together; cleaning and sub-fractionation were carried out using strong cation exchange (SCX) high performance liquid chromatography (HPLC). Briefly, sub fractions (1/3rd by volume, ~ 250 μL) of the pooled labelled peptides were resuspended in 40 μl of buffer A (5 mM Ammonium Formate, 30% (v/v) Acetonitrile, pH 2.3). Buffer B (500 mM Ammonium Formate, 30% (v/v) Acetonitrile, pH 2.3) and a SCX column (Zorbax 300, 150 × 2.1 mm, 5 μm, 300A, Agilent, USA) was used with following conditions: 0-5 mins at 100% A, 5-45 mins 100-30 % A, 45-50 mins 70-100 % B, 50-60 mins 100% B, 65-73 mins 100% A at a constant flow rate of 0.400 ml/min to prefractionate the peptide mixture.

**Figure 1:**
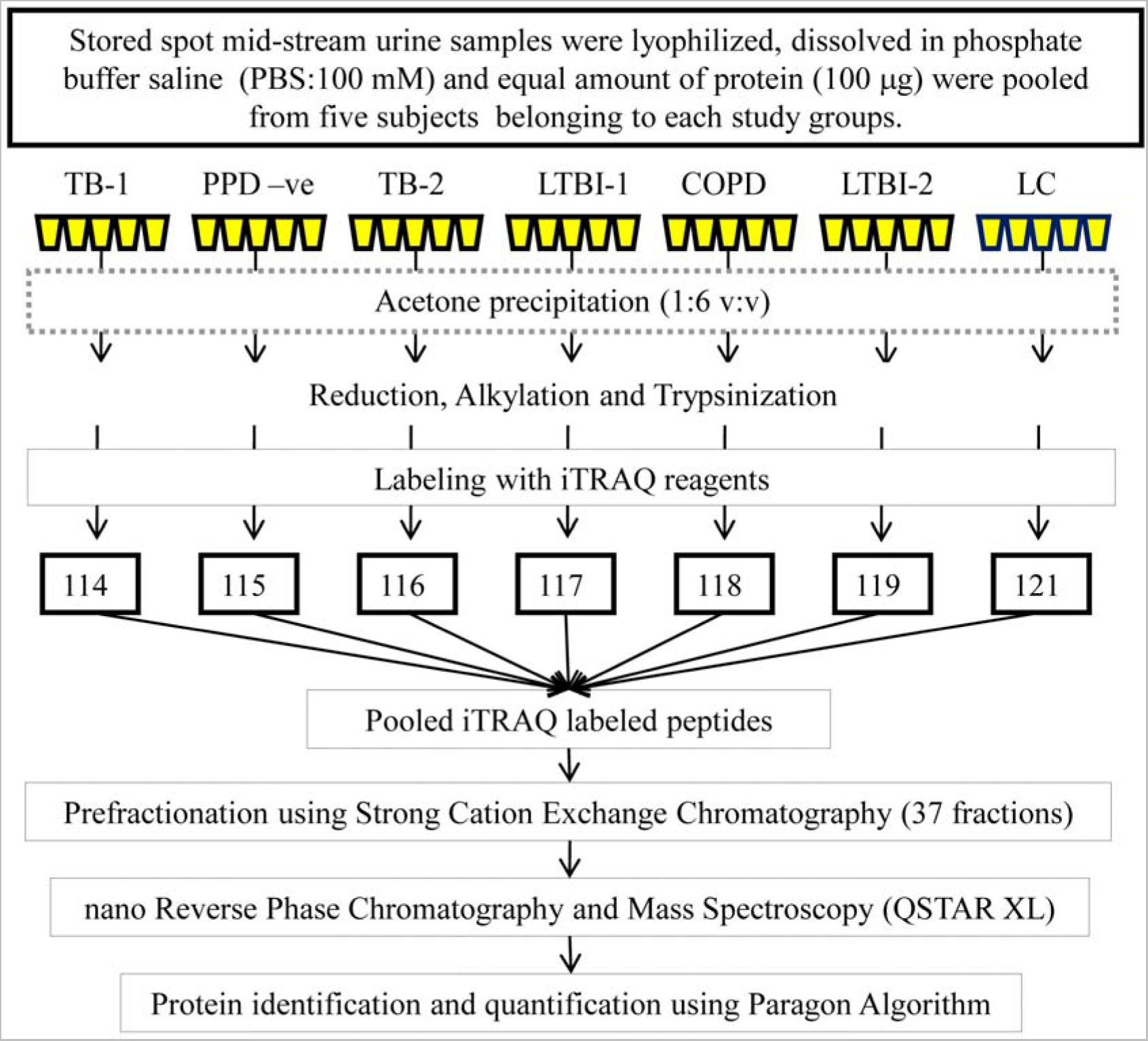
Urine biomarker discovery: a proteomics workflow for sample processing, iTRAQ labelling to mass spectrometry data analysis. Seven groups are used for the proteomics experiments with healthy and other disease controls. TB: active tuberculosis, PPD-ve: unexposed healthy subjects with –ve skin tuberculin test, LTBI: Latent tuberculosis infection, COPD: Chronic obstructive pulmonary disease and LC: lung cancer. 1 and 2 shows biological replicates.

A total of 37 peptide fractions were collected starting after a flow through of initial 5 minutes which are discarded. The fraction collected from 6^th^ to 8^th^ mins and 9^th^to 10^th^ mins were pooled to analyse as two fractions and named as fraction 1 and 2 respectively. From 11^th^ to 45^th^ mins every fraction was collected as fraction 3 to 37 separately for second dimension separation and mass spectrometry data acquisition. Each SCX fraction were dissolved in 15 μl of solvent A (0.1% formic acid in 3% Acetonitrile). These peptide fractions were separated on an online nano flow reverse phase HPLC system (Agilent 1100) and analyzed on a QSTAL XL mass spectrometer (Applied Biosystems, USA). Injection volume was set at 8 μl and Zorbax 300 SB-C18 (5x 0.3 mm) column (Agilent USA) was used for loading the peptides. Peptides were separated at 50 min linear gradient from 10% solvent A to 30% solvent B (0.1% formic acid in 90% acetonitrile) at an analytical flow rate of 250 nL/min using a Zorbax 300 SB-C18 (3.5 μM, 150 mm × 75 μm). The complete run time was 120 min for each SCX fraction including sample loading into enrichment column, elution from loading column, separation in reverse phase column and column reconditioning. Information-dependent data acquisition (IDA) was performed using Analyst software (Analyst QS 1.1) in positive ion mode at a spray voltage of 2.3 kV. Survey spectra were acquired with at a mass range from 450 to 1600 m/z. Molecules with charge state of 2 to 4 were selected for fragmentation and three most intense ions per cycle were isolated and fragmented at a mass range of 100 to 1500 m/z. Ions selected for fragmentation were dynamically excluded for 30 s after fragmentation. Ion tolerance was set at 100 ppm.

The MS/MS raw data of 37 fractions in .wiff file format (named 08122011U4 to U40) were analysed using ProteinPilot™ Software 4.0.8085 (Revision 148085 and source Applied Biosystems SCIEX, USA). These raw data files can be accessed by contacting the corresponding author by email. Raw data files were converted to searchable MS/MS peak lists automatically without merging the putatively like spectra. In built Paragon™ Algorithm (4.0.0.0, 148083) present in Protein Pilot was employed to identify peptide sequence tags, perform data base matching for protein identification, protein grouping to remove redundant hits and comparative quantification. The *Homo sapiens* protein database (UniProt/Swiss-Prot version, released March 14, 2011 with 61,113 proteins) was used for all searches. Trypsin was used to generate peptides with a default probability at 0.00005, misscleavage factor of 0.75 with a cleavage rule after Arg with probability of 0.9 (exception before proline at 0.15, Aspartic Acid of 0.70, Glu of 0.85) and cleavage rule after Lys with probability of 0.80 (exception before Pro at 0.034, Lys at 0.7, Asp of 0.50, Glu of 0.60). Cys alkylation was carried out by methyl methanethiosulfonate (MMTS) and iTRAQ labelling at N-terminal and Lys side group of peptides were selected as fixed modifications. Special factors and identification focus were set as was selected by Gel-based ID and biological modifications respectively. Mass tolerance for precursor ions was set as fraglet_correction at 1, quant_threshold 9, mass tolerance for fragment ions for auto calibration msmstolsd ratio was 4 and mstolsdratio 4 dalton. Both MS and MS/MS tolerance values were set at0.2 dalton. MS and MS/MS standard deviation values were set at 0.0125 dalton and 0.03 dalton respectively. Data was normalized for loading error by bias corrections calculated using ProteinPilot. All protein identification was based on reported commonly acceptable 95% confidence and is determined by ProteinPilot (threshold score as ProtScore > 1.3). A further requirement was a protein p-value, which ensured protein identification and quantification was based on peptide or spectra hits. False discovery rate (FDR) at protein levels was calculated using the standard inbuilt algorithm available with Protein Pilot. Quantification of proteins was carried out by taking the average ratio including correction for both experimental bias and background. The reported average ratio is inversely weighted by the % error calculated for each distinct peptides identified. As per the sample availability for each study groups biological replicates were included. A representative mass spectrum of peptides that were used to identify and quantify a representative protein (zinc alpha-2-glycoprotein) is presented in Figure 2.

**Figure 2:**
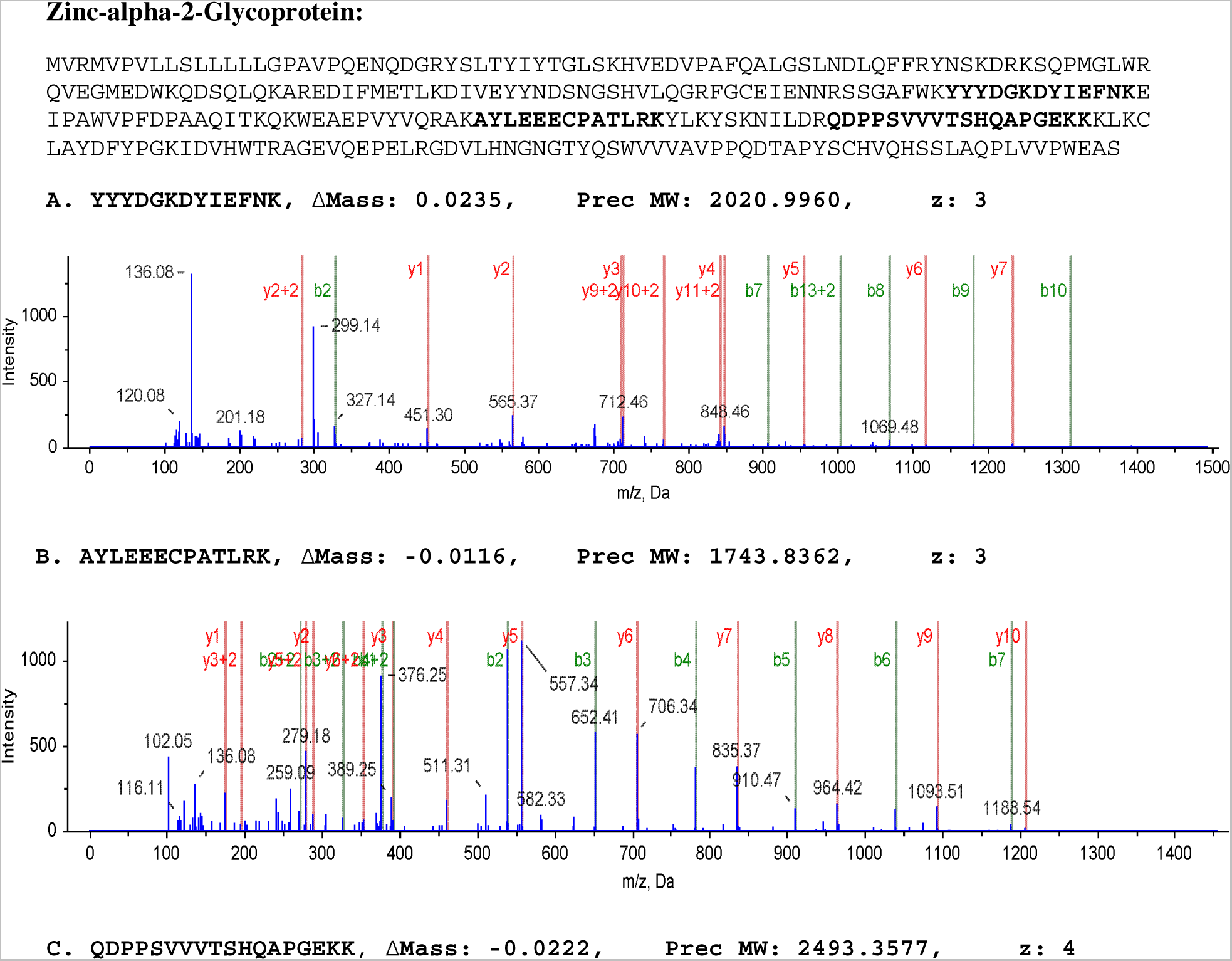
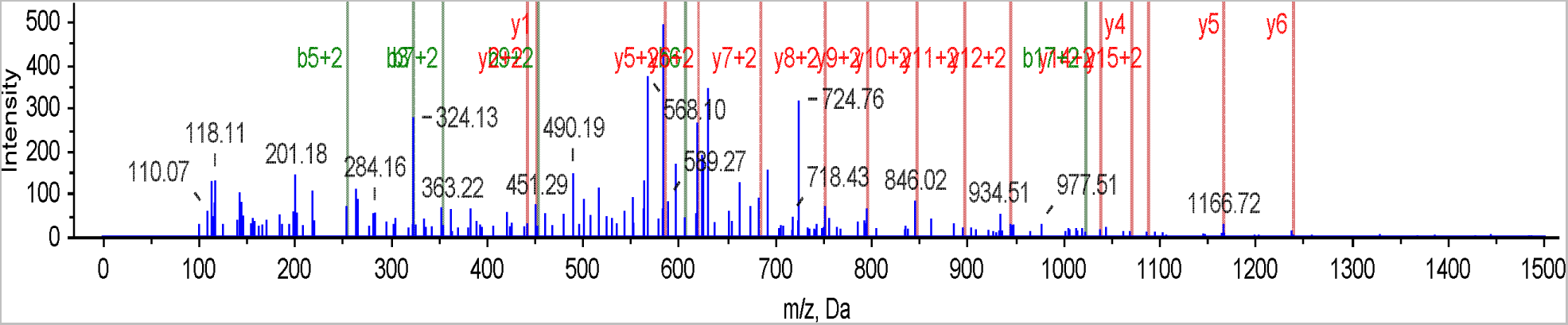
Representative mass spectra data of one of the marker protein Zinc-alpha-2 glycoprotein showing contributing three peptides (out of eight) used for identification and quantification. ΔMass: mass difference, Prec MW: predicted mass, z: charge

### Construction of functionally associated urine protein network of healthy and TB

TB urine proteome sub-network was constructed with 83 differentially expressed proteins observed in at least one TB group were considered in this study. We created a union dataset of urine proteome reported in three studies (12–14). In total there were 4387 genes with an overlapping set of 2477 genes reported in these studies. We generated a non-redundant set of 2563 genes to build the functionally associated healthy urine protein network. While constructing the functional association protein sub-network for urine proteome in TB we set the path length of two, so that two nodes can be connected if there is one intermediate protein. A path length of 2 here refers the requisite of two edges to connect two seeds; hence there could be only one intermediate protein between them. The edges in all protein-protein interaction network (PPI) networks were treated as undirected. Concurrently, we constructed the network of healthy urine proteome from the proteomics data collected from different studies using path length of one. PPI databases that were used for network construction includes: BIND, DIP, PDZBase, IntAct, HPRD, InnateDB, BIOCARTA, MINT, MIPS, KEGG and BioGRID. Only curated PPI databases were considered to build PPI network so as to minimise false positives. To further ensure the quality of the interactions in TB network, we set the choice that the protein-protein interaction must have reported in at least two articles. We queried the healthy urine proteome network with the TB urine proteome network that is inferred from this study. Henceforth, we refer to healthy urine proteome network as reference and TB urine proteome network as query network. The topological parameters of reference and query PPI networks were analysed using NetworkAnalyzer (15) plugin of Cytoscape version 2.8.3 (16). The details of the statistical measures used for network comparison can be found in greater detail from the published articles of NeAT (Network Analysis Tool) tool (17).

### Bioinformatics Analysis

Over-representation analysis (ORA) was performed by considering the whole human genome as reference set using Bingo 2.4 plug-in (18). For multiple hypothesis testing, we used Benjamini and Hochberg based p-value correction method for FDR correction, although less stringent than the alternate Bonferroni family wise error rate (FWER) method. Enrichment map (19) and WorldCloud plug-ins (20) were used for visualization of enrichment analysis. We constructed the functionally associated network of proteins for the urine proteome from healthy and TB data using X2K (21). Network comparison analysis was carried out using NeAT (Network Analysis Tool) (17). The ClusterONE plugin 0.93 (22) was used to identify functional modules in the TB urine proteome network.

### Statistical Analysis

As the study attempts to identify putative biomarkers, we identified the proteins showing significant differential expression (at least 2-fold up or down) and for this purpose all protein iTRAQ ratios were transformed to base 2 logarithm values. In base 2 logarithm space, a 2 fold alteration in levels is reported as −1 and 1 for down-regulated and up-regulated changes with the p-value <0.05, respectively and identified as hits.

## Results

In the iTRAQ experiment, we identified a total of 131 proteins at 95% significance (ProtScore > 1.3) and false discovery rate (FDR) less than 1% at protein level. Proteins showing differential expressions in one of the TB groups with respect to healthy subjects are presented in Figure 3. We observed 13 proteins showing two-fold up or down regulation in at least one of the two active TB groups at p ≤0.05 (Table 2, Figure 3). It is to be noted that after correcting the p-value for multiple hypothesis testing problem, p-value remained insignificant. Two out of 13 proteins namely Q8WZ75 (Roundabout homolog 4) and P15309-2 (Isoform 2 of Prostatic acid phosphatase) were down-regulated in active TB groups. Some of the proteins in the up-regulated list include Q53H26 (Transferrin), Q4W597 (Osteopontin), B3KRN9 (Uromodulin), Q96CZ9 (Cadherin 11), P04279 (Semenogelin-1), P25311 (Zinc-alpha-2-glycoprotein), Q6Q3G8 (Lysosomal-associated membrane protein 2) and P02750 (Leucine-rich alpha-2-glycoprotein). We identified in total 11 proteins that show significant alteration in abundance in LTBI (Supplementary Table S1). We observed 10 proteins, most of them consistently showing up-regulation in both the LTBI groups and one protein Q6P5S3 (uncharacterized protein) getting down-regulated.

**Figure 3:**
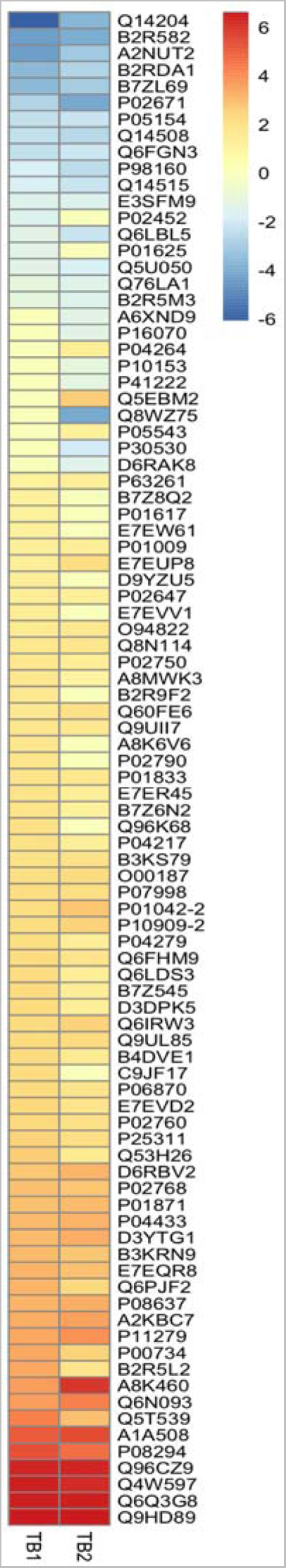
List of deregulated urine proteins observed in active tuberculosis (TB) patient groups with respect to unexposed healthy subjects with –ve skin test results (healthy).

**Table 2:** Differential expression of proteins identified in two TB groups.

| **Uniprot**<br>**Accession ID** | **Protein Description** | **TB**<br>**GP1** | **p-val** | **TB**<br>**GP2** | **p-val** |
| --- | --- | --- | --- | --- | --- |
| **P25311** | Zinc-alpha-2-glycoprotein | 2.51 | 0.001 | - | - |
| **P04279** | Semenogelin-1 | 2.15 | 0.058 | - | - |
| **P02750** | Leucine-rich alpha-2-glycoprotein | 1.62 | 0.04 | - | - |
| **Q6Q3G8** | Lysosomal-associated membrane protein 2 (UNIPROT replaced ID: P13473) | - | - | 6.46 | 0.02 |
| **Q96CZ9** | Cadherin 11 | - | - | 6.34 | 0.02 |
| **Q4W597** | osteopontin (UNIPROT replaced ID P10451) | - | - | 6.23 | 0.05 |
| **E7EQR8** | Uncharacterized protein GN=YIPF3 | 3.10 | 0.057 | 2.92 | 0.03 |
| **B3KRN9** | Uromodulin (UNIPROT replaced ID: P07911) | 3.06 | 0.014 | 2.76 | 0.03 |
| **P01833** | Polymeric immunoglobulin receptor | 1.82 | 0.016 | 1.63 | 0.00 |
| **Q53H26** | Transferrin | 2.60 | 0.003 | 1.49 | 0.01 |
| **E7ER45** | Uncharacterized protein GN=MGAM | - | - | 1.25 | 0.05 |
| **P15309-2** | Isoform 2 of Prostatic acid phosphatase | - | - | -2.03 | 0.02 |
| **Q8WZ75** | Roundabout homolog 4 | - | - | -4.15 | 0.01 |

TB-GP1 refers to proteins identified in TB group 1 and TB-GP2 refers to proteins identified in TB group 2. “-” refers to proteins undetected in the groups. In case of uncharacterised proteins, wherever applicable, the gene name is given by value of “GN”.

### Comparison of differential pattern across related pulmonary diseases

Cross-verification of the differential expression pattern in the related pulmonary diseases such as COPD and lung cancer was carried out. Twelve proteins showed significant alteration (up-regulation) in COPD subjects (Supplementary Table S2), out of which only protein expansion/expression α1-Microglobulin/Bikunin Precursor (AMBP) was found to be specifically up-regulated in COPD. Lung cancer subjects showed the altered expression of 15 proteins (Supplementary Table S2), out of which 10 proteins were specific to it. Out of total 27 proteins reported as differentially excreted in urine (Table 2, Supplementary Tables S2 and S3), 16 proteins were observed in one of the diseases (TB or LTBI or COPD or LC), single protein in two diseases (LTBI and COPD), seven proteins in three diseases [(LTBI, COPD and LC) or (LTBI, COPD and TB) or (COPD, TB and LC)] and three proteins in all four disease groups. Out of the 27 proteins, 4 are specific to TB: Q8WZ75 (Roundabout homolog 4), P15309-2 (Isoform 2 of Prostatic acid phosphatase), P04279 (Semenogelin-1), P02750 (Leucine-rich alpha-2-glycoprotein), 1 for COPD: P02760 (Protein AMBP), 1 for LTBI: Q6P5S3(Putative uncharacterized protein), 10 proteins for lung cancer: P01034 (Cystatin-C), Q7Z3B1 (Neuronal growth regulator 1), Q9UL83 (Myosin-reactive immunoglobulin light chain variable region (Fragment), B7Z545 (cDNA FLJ60068, highly similar to Inter-alpha-trypsin inhibitor heavy chain H4), B2R888 (Monocyte differentiation antigenCD14), O00187 (Mannan-binding lectin serine protease 2, P16070 (CD44 antigen), P06870 (Kallikrein-1), P63261 (Actin) and P01008 (Antithrombin-III). It is to be noted that all 10 altered proteins in lung cancer were down-regulated. The three proteins that are found to be up-regulated in all four disease groups include, E7ER45 (Uncharacterized protein), Q4W597 (Osteopontin) and P13473 (Lysosome-associated membrane glycoprotein 2).

### Comparison of TB urine network and healthy urine network

In a PPI, the nodes represent the proteins and the edges represent the interactions. The resultant reference network constituted 2563 nodes and 4807 edges, while the query graph contained 159 nodes and 401 edges (Table 3). As most of biological networks, we observe that degree distribution in the healthy urine proteome network following a power law [P (k)~ak^b^] (a = 715.05; b = −1.551; correlation =0.936; R^2^ = 0.867). It is observed that the number of proteins declines very quickly until about 15 degrees, indicating that there are very few proteins containing more than 15 interaction partners in the network. Power-law agreement is also observed for TB network (a = 35.380; b = −1.018; correlation=0.439; R^2^ = 0.564). The intersection between healthy and TB urine networks is minimal having 105 nodes and 139 edges in common, which represents less than 5% (Jaccard coefficient of similarity =0.027at p<10^−287^) of the union (i.e. healthy and TB urine proteome) (Figure 4, Table 4). However, the number of expected edges is still significantly very low (0.56).While our interest is to identify putative biomarkers, we observed that difference in the two networks was composed of 54 nodes and 262 edges (Figure 5) (Supplementary Table S4).

**Figure 4:**
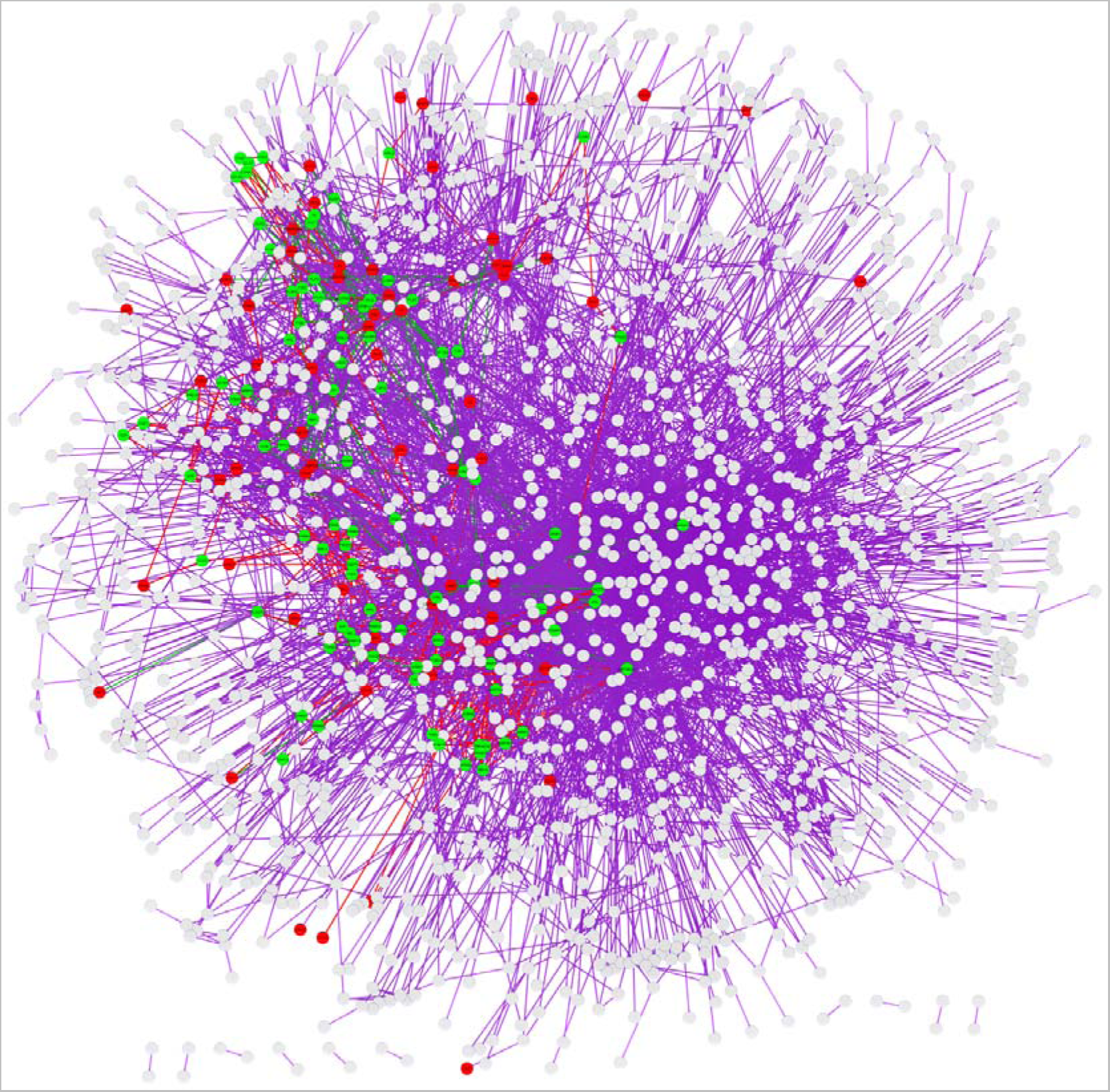
Network comparison of healthy and TB urine proteome network. The union graph computed by NeAT tool for the functional associated network of TB and healthy urine proteome. The colour code reflects the status of the arcs present at the intersection of query (Q) or Reference (R) or in one graph only. Arcs found at the intersection between graphs R and Q are colored green. Arcs found in graph R but not in graph Q are colored violet. Arcs found in Q graph but not in graph R are coloured red.

**Figure 5:**
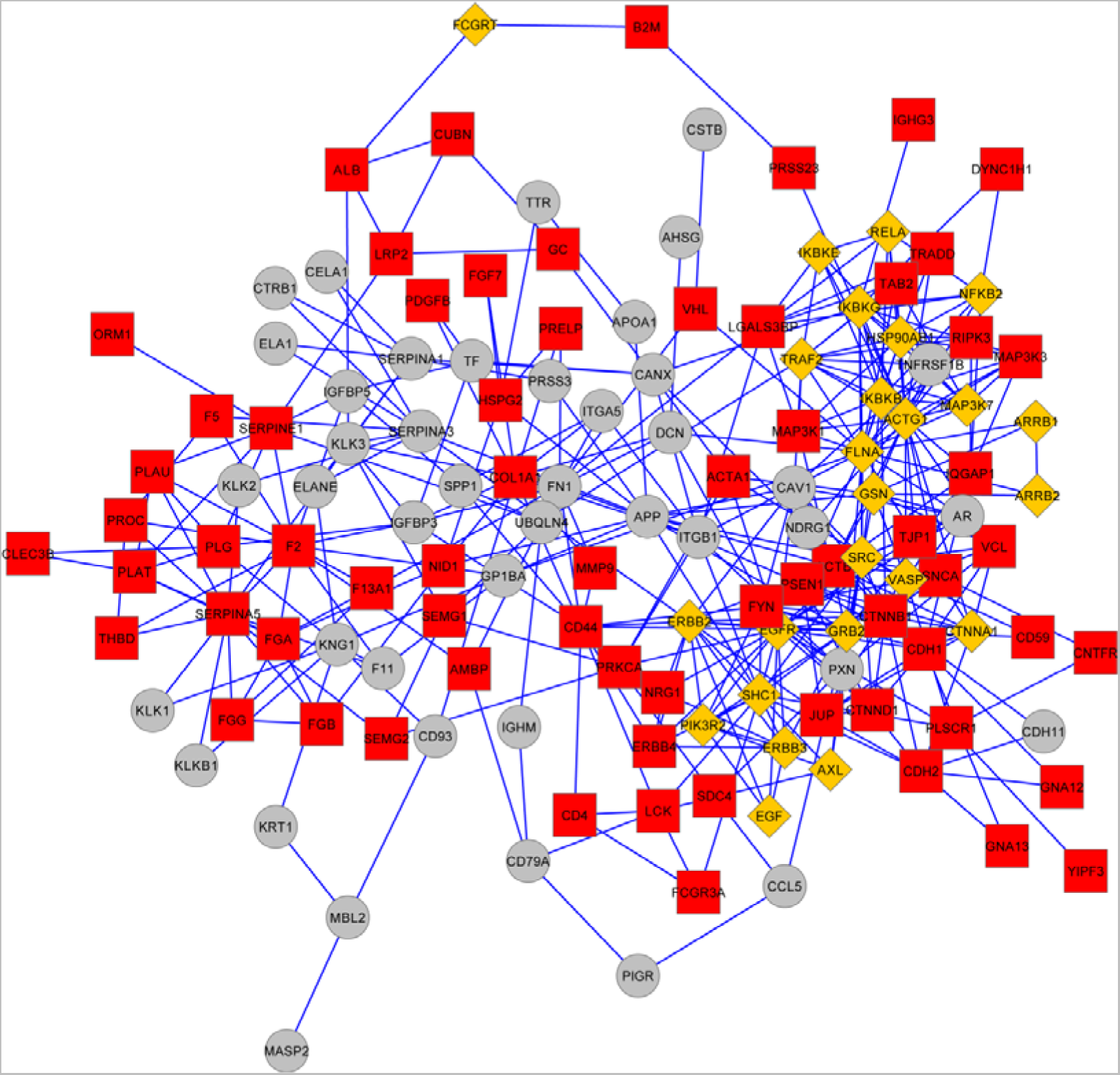
Identification of protein-protein interaction (PPI) clusters in TB urine proteome network. The network shown here with 54 nodes and 262 edges is clustered by ClusterONE method. The nodes of the network are colored according to the number of clusters they participate in. Nodes that correspond to a single cluster are coloured red; nodes belonging to multiple clusters are colored yellow. Outliers that do not belong to any of the cluster are colored grey.

**Table 3:** Topological characteristics of networks compared in this study.

| Graph | Nodes | Arcs | Description |
| --- | --- | --- | --- |
| **R** | 2563 | 4807 | Reference graph |
| **Q** | 159 | 401 | Query graph |
| **Q∪R** | 2617 | 5069 | Union |
| **Q∩R** | 105 | 139 | Intersection |
| **Q!R** | 54 | 262 | Query not reference |
| **R!Q** | 2458 | 4668 | Reference not query |

Q – Query graph (TB urine proteome network) R –Reference graph (Healthy urine proteome network)

**Table 4:** Network Comparison Statistics.

| Symbol | Value | Description | Formula |
| --- | --- | --- | --- |
| **N** | 2617 | Nodes in the union |  |
| **M** | 3423036 | Max number of arcs in the union | $M = N*(N-1)/2$ |
| **E(Q^R)** | 0.56 | Expected arcs in the intersection | $E(Q^R) = Q*R/M$ |
| **Q^R** | 139 | Observed arcs in the intersection |  |
| **perc_Q** | 34.66 | Percentage of query arcs | $perc\_Q = 100*Q^R/Q$ |
| **perc_R** | 2.89 | Percentage of reference arcs | $perc\_R = 100*Q^R/R$ |
| **Jac_sim** | 0.0274 | Jaccard coefficient of similarity | $Jac\_sim = Q^R / (Q \vee R)$ (intersection size divided by union) |
| **Pval** | 3.00E-287 | P-value of the intersection | $Pval = P(X \geq Q^R)$ |

### Functional module identification

Clustering analysis of PPI network helps in the identification of functional modules of genes that collectively mediate the molecular functions. Clustering of TB PPI network resulted in 19 clusters, with a size ranging from 3 to 17 members (Figure 6). However, we could observe only four statistically significant clusters (p-value <0.05). The top ranking cluster that involved 17 genes revealed the over representation of several important functions that were not visible before the network expansion. These included the molecular functions namely: positive regulation of intracellular protein kinase cascade (specifically I-kappaB kinase/NF-kappaB cascade), receptor signalling protein serine/threonine kinase activity, signal transmission via phosphorylation event, MAP kinase activity, positive regulation of apoptosis and negative regulation of protein catabolic process. The third ranked cluster is found to collectively mediate transmembrane receptor protein tyrosine kinase signalling pathway, positive regulation of phosphoinositide 3-kinase cascade, negative regulation of apoptosis etc.

**Figure 6:**
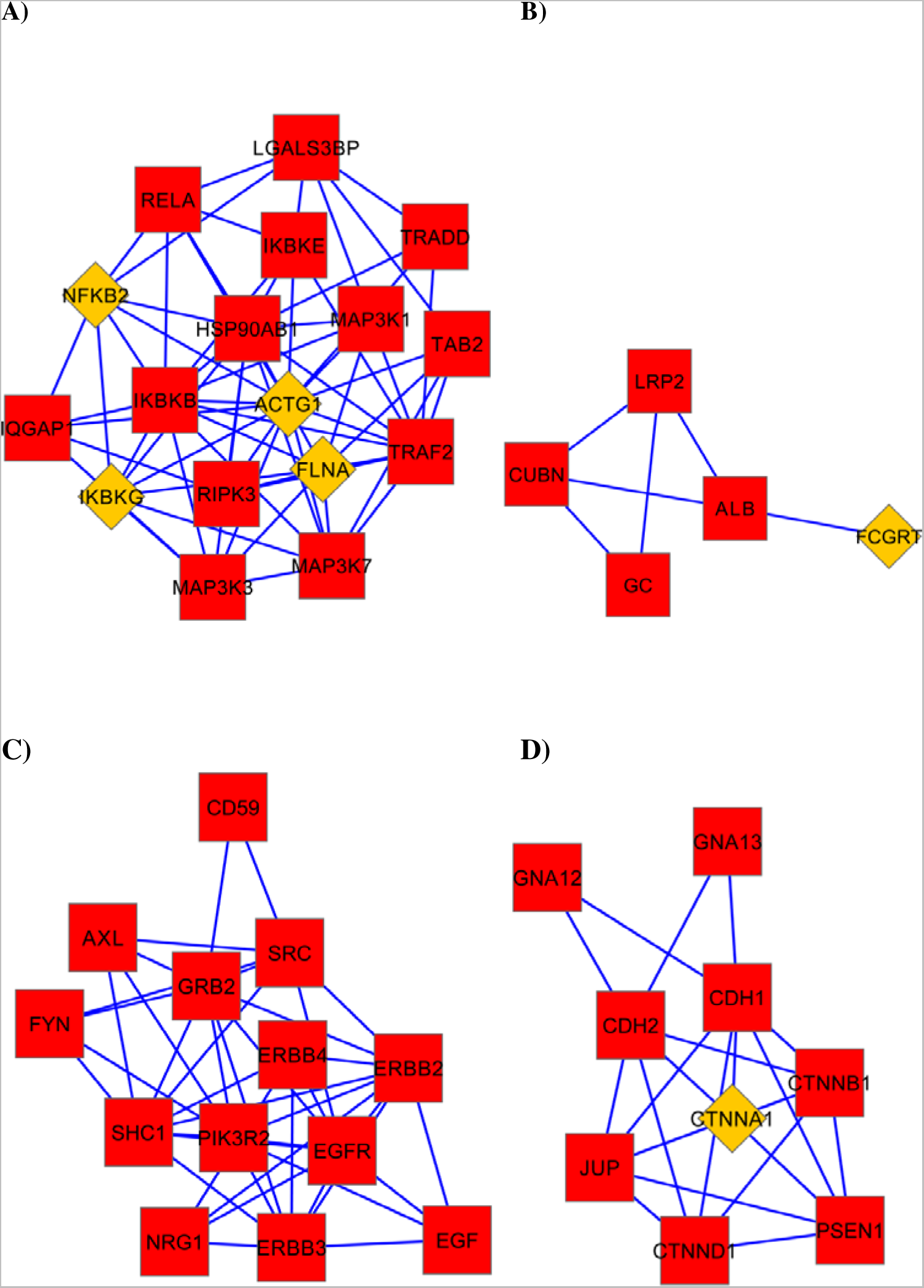
Extraction of significant protein-protein interaction (PPI) cluster from TB urine proteome network. Top ranked clusters according to their p-values are shown here. A) Cluster with 17 members (density 0.478), B) Cluster with 5 members (density 0. 6), (C) Cluster with 13 members (density 0.526), D) Cluster with 9 members (density 0.611). Nodes that correspond to a single cluster are colored red; nodes belonging to multiple clusters are colored yellow.

### Over Representation Analysis (ORA)

The list of proteins showing similar fold changes in both the TB groups irrespective of their p-values is presented in Supplementary Table S3. The proteins for which the gene names were mapped considered for over representation analysis (ORA) to determine the predominant biological functions represented by the subset of TB urine proteome identified in this study. About 70% of the proteins identified in urine proteome of TB patients had an extracellular origin, while remaining belong to membrane-bound vesicle, stored secretory granule etc. The up-regulated genes were found to over-represent molecular functions that includes acute inflammatory response, regulation of signalling pathway, calcium ion binding, localization, homeostatic process, signal transduction, regulation of signalling process, immune system process, response to stress, serine-endopeptidase inhibitor activity, biological adhesion, cell adhesion, transport, lipid binding, platelet-derived growth factor binding, calcium oxalate binding etc (Figure 7 and Supplementary Table S4). The down-regulated genes were over-represented in the molecular functions some of which includes endopeptidase inhibitor activity, regulation of cell adhesion, beta-N-acetylgalactosaminidase activity, prostaglandin-D synthase activity, antigen processing and presentation of exogenous peptide antigen via MHC class Ib protease binding, response to wounding etc (Supplementary Table S5). It is noteworthy to mention that some of the biological functions like regulation of apoptosis (as in ORA of functional modules), endopeptidase inhibitor showed variation in both directions.

**Figure 7:**
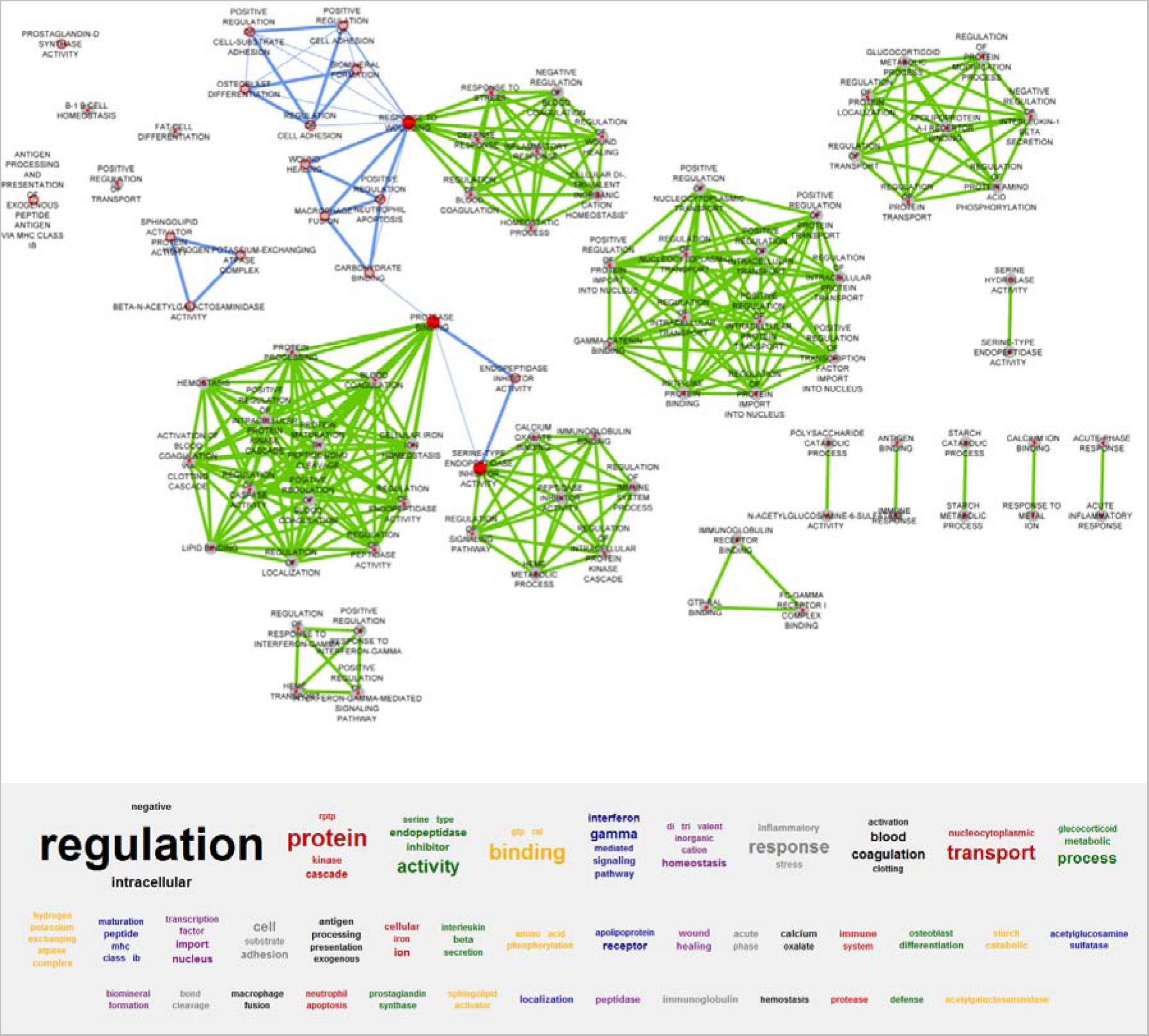
Gene set enrichment analysis of TB urine proteome. Enrichment Map, a Cytoscape plugin was used for visualizing the enrichment results produced by Bingo. WordCloud plugin was used to summarize the biological terms. Nodes represent enriched gene sets, which are grouped and annotated by their similarity according to related gene sets. Node size corresponds to the total number of genes within each gene set. Edge thickness corresponds to the number of shared genes connected gene sets. Up-regulated gene set are shown in green while down-regulated genes are show in blue.

## Discussion

The abnormally expressed proteins in diseased conditions are detected in urine. We try to identify the interacting partner, as proteins might not function in isolation in cellular system. By PPI, we identify those interacting partners that mediate certain functions in the system before their excretion in urine. With the existing urine proteome data from healthy and current experiment determined proteome data of TB samples, we construct the sub-network for comparison. Although the network doesn’t have quantitative information, network comparison derives the difference network based on statistical significance. PPI network and over-representation analysis may aid the understanding the functional roles of the proteins identified.

Although the spot urine samples show somewhat higher variability as compared to morning sample, it offers two advantages: as processing time in bladder is less, it is susceptible to minimum proteolytic cleavage and secondly it is practically convenient for collection (23). Urine processing for proteomics studies has been discussed in multiple reports. Depletion strategies to remove high abundant proteins like albumins and immunolglobulins have shown to decrease the overall complexity of urine samples (24). However a mixed opinion also exist that removal of these mostly hydrophobic proteins could lead to removal of other low abundant interacting proteins which may aid disease classification. This implies a trade-off between dynamic range and complexity and therefore we did not undertake any high abundant protein depletion from samples in this study. As TB is a complex disease, resultant immune response involves a complex network of proteins. We identify several proteins in this study that could possibly play a crucial role in the pathogenesis.

Although we identified several proteins that showed significant fold change in TB subjects, our primary aim was to identify the most specific markers of TB while comparing the other groups considered in this study. The study brings out an important outcome of deregulated proteins specific to each disease condition. For example, we notice specific up-regulation of leucine rich alpha-2-glycoprotein (LRG), a secretary protein that was differentially expressed only in urine of TB samples. Similar findings for LRG have been reported in serum and pericardial effusion of TB patients (25). With abnormal expression of LRG in TB and its proven clinical importance in other diseases like appendicitis (26), we anticipate the potential application of LRG as a marker, which however needs further validation.

Robo-4 is well studied as a tumor-specific endothelial marker owing to its discriminate expression pattern between tumour and healthy cells (27). Slit/Robo signalling pathway plays a crucial role in regulating cell proliferation and cell motility which are often deregulated in cancer, and hence, one of the key component Robo-4 has been proposed as potential therapeutic target, for ex in Hematopoietic stem cells (HSC) transplantation therapy (28).The down-regulation of Robo-4 in TB disease as observed in this study denotes its potential application as a specific marker for TB and it will be an important study to unveil the functional role of Robo-4 in TB pathogenesis.

Another putative marker, Semenogelin-I, a prostrate specific secretary protein reported in human urine (29) showed significant up-regulation in active TB samples. Ectopic release of semenogelins are also been reported from lung cells in different disease conditions and fragmented peptides of it have shown antibacterial activity (30). This suggests a possible secretion of semenogelin from Mtb infected lung tissues that may perhaps protect the host by its antibacterial activity that needs further validation.

Lysosomal associated membrane protein 2 (LAMP-2) is a marker found in both late endosome and lysosome membrane (31), that shows quantitative variation in phagosome of Mtb infected and healthy cells (32). Although the function of LAMP-2 is obscure, it is believed to mediate transport of cellular materials or digestive enzymes to the lysosome through autophagosome. In addition it also plays a critical role in the fusion of autophagic vacuole with lysosomes. Phagosome maturation is affected in TB and this could possibly stipulate increased expression of LAMP-2 in both latent and active TB cases to increase lysosome fusion. However Mtb has evolved other successful strategies like iron accumulation, pH alteration and others to inhibit fusion with lysosome to survive in the host. As observed in our study LAMP-2 expression is significantly altered in all study groups and hence its specificity for TB is uncertain.

Cadherins plays an integral role in cell adhesion in a calcium dependent manner. In this study we observed the increase in abundance of cadherins in active TB and COPD, while it is down-regulated in LTBI. These results suggested that the loss of E-cadherin-mediated adhesion might provide a spatial cue for the generation of mature, migratory dendritic cells (DCs) but without the ability to induce T cell immunity. Earlier reports also showed that E-cadherin plays as a first cellular receptor for the internalization of *Listeria monocytogenes* (33). Additionally it also increases cell permeability that may lead to the release of cadherin molecules from the infected tissues to circulatory fluids thereby increasing cadherin concentration in urine of infected patients (34). Abnormal expression levels of E-cadherin are reported in other disease conditions like non-small-cell lung carcinoma (NSCLC), diabetic nephropathy and bladder transitional cell carcinoma and have been investigated as potential biomarkers with prognostic ability (35–37).

Another putative marker identified in this study, osteopontin functions as a Th_1_ cytokine and macrophage chemoattractant that plays crucial role in TB pathogenesis. Possible role of osteopontin in TB pathogenesis by inducing IL-12-mediated type-1 T helper cell response in *in vitro* experiments has also been suggested previously (38, 39). Furthermore, increased expression of osteopontin gene has been reported in Mtb infected primary human alveolar macrophages (40). The role of osteopontin in granuloma formation which is common in TB has also been investigated in the same study. Increased plasma osteopontin concentration is observed in TB and our finding from urine corroborates it. Osteopontin also shows significant alteration with therapeutic interventions which make it as an ideal prognostic marker for TB (38, 39).

We noticed up regulation of uromodulin in both active and latent TB study groups and as well as COPD subjects, however in LC it is down-regulated. Uromodulin or Tamm-Horsfall glycoprotein (THP) is found in high abundance in urine of healthy subjects yet its function remains unclear (41). Some of the known functions of uromodulin include antigen induced proliferation of human lymphocytes in vitro: potent inhibitor of interleukin 1 (IL-1)-induced thymocyte proliferation and inducer for IL-2 induced thymocyte proliferation (42). Previous studies have suggested antibacterial activities of uromodulin especially in urinary tract infections (43). Speculative anti-bacterial function of this protein in TB is very unclear at this stage and needs a detailed study.

It is well known that iron (Fe) plays a critical role in the growth and sustenance of both host and Mtb. Host plays an evolutionary strategy to limit the use of iron by chelation of extracellular iron by host proteins such as transferrin, lactoferrin and intracellular ferritin. Intracellular growth of Mtb (virulent Erdman strain) in human monocyte derived macrophages showed Fe acquisition from extra-cellular transferrin (44). In our study we observed increased abundance of transferrin in urine of both latent and active TB patients. This is suggestive of a host defence role to compete and there by deprive the intracellular uptake of Fe in Mtb. Some of the intracellular pathogens including Mtb inhibit respiratory burst using tartrate-resistant non-specific acid phosphatases (ACPs) and enhance their survival in phagocytic cells. Decreased abundance of phosphatases in urine of TB patients explains larger utilization of these molecules inside the infected tissues. However the mechanism of decreased release of prostatic acid phosphates from its origin i.e. prostatic glands needs further investigation.

Increased expression of zinc α2-glycoprotein (ZAG) in urine of active TB, latent TB and COPD patients is observed which may be to satisfy the increased demand of infected tissues for fatty acids. ZAG stimulates lipid degradation in adipocytes, and acts as a lipid-mobilizing factor and causes extensive fat losses in certain cancer conditions (45). Its role in additional binding ability to polyunsaturated fatty acids for transportation has also been studied. Importance of ZAG as a protective serum biomarker candidate has been investigated for prostate cancer (46).

Apolipoprotein A-I (Apo A-I) is one of the well-studied acute phase response protein which gets down regulated during infection. ApoA-I is known for its anti-atherogenic, antioxidant, anti-inflammatory properties and anti-bacterial activity by promoting complement-mediated killing of gram-negative bacteria. ApoA-I is the major constituent of high density lipoproteins (HDL) that plays the functions of: promoting cellular cholesterol efflux, acting as a cofactor for the lecithin cholesterol acyltransferase (LCAT) and forming mature HDL (47). HDL associated Apo A-I works as negative acute phase protein and its level goes to 0.25 fold in certain acute phase response (48). Similarly, in this study we observed a decrease of more than 0.5 fold in TB patients. Decrease in Apo-A-1 positively regulate tumour necrosis factor-α (TNF-α) and interleukin −1β, the essential components for controlling host defence in both active and latent TB pathogenesis (48). With the up-regulation of these proteins in TB, it is not clear if ApoA-1 might play a similar role against Mtb. Further studies investigating the role of ApoA-1 in mediating anti-mycobacterial activity may be helpful.

We observed the deregulation of proteins identified from the difference sub-network in other conditions and their functions are found to be related to assisting or resisting the infections. Serpin peptidase inhibitor plays important role in protein metabolism and its function involves as principal inhibitor of tissue plasminogen activator (tPA), urokinase and inhibits fibrinolysis. Higher concentration of this gene product is associated with thrombophilia and up-regulated Serpin peptidase inhibitor gene is reported in TB meningitis (49). Another protein we identified from network comparison is catenin which is also reported to be involved in TB pathogenesis. Studies have shown that BCG infection down regulates β-catenin (adherens junction protein) expression and thereby increases permeability across confluent mesothelial monolayer. Inverse regulation of the TLR/NF-κB and the Wnt/β-catenin pathway in BCG infected murine model is also reported. The study demonstrated the involvement of Wnt/β-catenin pathway in tissue homeostasis that gets switched off under proinflammatory conditions (50).

It is well known that host cell signalling pathways are efficiently exploited by Mtb for its survival inside the host. Phagolysosome fusion via phosphoinositide 3-kinase (PI-3K) signalling pathway is hampered in presence of LAM secreted from virulent Mtb (51). Reports also show addition of Vitamin D3 reverse the phenotype of phagosomes and promote phagolysosome fusion by PI-3-K pathway (52). Although these proteins were not identified in the proteomics experiment these has been extracted from the network analysis. Mannan-binding lectin (MBL) is reported to be present in serum and play an important role in host defence against pathogens by contributing critical role in the innate immune system. It has an ability to bind ligands, for example, specific carbohydrate moieties like LAM present on Mtb cell wall and activates the classical pathway of complement activation via an activation of the Clr2Cls2 complex (53). In Indian population, researchers have showed that lower expression of mannose binding protein (MBP) genes and different allelic variants make them susceptible to different infections including pulmonary TB in different age groups (54). We do expect lower MBP concentration in the serum and urine of TB patients and could be useful as an important marker for TB.

Urine proteins identified from samples of active TB patients covered 5% of total healthy dataset collated from three different studies (12–14). Presumably, the list of urine proteins might be still higher, yet it is suitable in this case to compare the coverage and understand the difference in the proteomic content with that of diseased. By mapping the interacting partners of the deregulated proteins of the TB urine proteome, we determined those possibly deregulated proteins, which we perhaps missed to capture in the experiment. While comparing the functionally associated network of healthy and TB, we observed the sub-network formed by 54 proteins with 262 interactions specific to TB. We hypothesize that the difference network, could array a list of deregulated proteins and enrich the perturbed functions of TB pathogenesis that is explained by the experimentally identified proteins. However it is to be noted that we did not consider the intermediate partners of the reference network, which might result in disproportionately large reference network. We expanded the PPI network of proteins identified in TB patient urine samples, basically to observe the putative coverage of proteome that could have been possibly identified in this study but may be missed due to certain limitations in adopted experimental procedures and mass spectrometer. We bridged the missing links by fetching them from the repertoires of experimentally verified protein-protein interactions.

The over-representation of molecular functions of the experimentally identified proteins is populated with the more members after network expansion. For example, before the network expansion, ORA analysis revealed the molecular function GO term: calcium binding was represented by 9 gene set (CDH1, CDH11, CDH2, EGF, F2GSN, MASP2, PRSS3, and SPARCL1) most of which are up-regulated. After network expansion we observed the enrichment of 10 genes (CANX, CD93, CUBN, LRP2, NID1, PLG, PLSCR1, PROCS, NCA, and THBD) to the GO term, which we hypothesize that could be involved in TB pathogenesis and might also be differentially expressed although not identified in this study. In support of our hypothesis, we found literature relating some of these genes to infection related pathways.

Activation of ER stress pathway leads to up regulation of associating molecules like calnexin in the granulomas isolated from lungs of TB patients (55). CD93 scavenges the apoptotic cells either by influencing defence collagen or by CR1-mediated phagocytosis. CD93 work in association with moesin and involve the phosphatidylinositol 4,5-bisphosphate (PIP-2) to execute its function (56). Intriguingly, Mtb also encodes a close homolog of nidogen, PknD, which has a sensor domain, capable of forming a NHL-domain β propeller (57). The sensor domain of Mtb PknD, has been suggested to facilitate the adhesion of Mtb to the micro vascular endothelium of the CNS (58). In certain bacterial invasion like *Enterococcus faecium*, nidogen is found to work as a potential ligand however its role in Mtb interaction needs further investigation (59). Another interesting protein identified in the difference network is plasminogen, which when converted to its active form i.e. plasmin dissolves fibrin clots by degrading extracellular matrices directly or indirectly. It has been well documented that different types of bacteria interact with the plasminogen system by different mechanism to involve in bacterial pathogenesis. Reports also show that Mtb possesses plasminogen binding as well as activating molecules and allow them to break down the fibrin meshes resulting from inflammatory processes and blood basement membranes to facilitate host invasion and systemic dissemination (60). Phospholipid scramblase 1 plays important role in regulation of lipid accumulation and found to be significantly up-regulated in human TB granulomas (61). Thrombomodulin is a glycoprotein receptor plays major role in activated protein C (APC) generation that mediates both anticoagulant and anti-inflammatory properties in lungs. Loss of function as mutation of thrombomodulin gene leads to the enhanced rate of Mtb mortality by an uncontrolled inflammation response in mice (62). Protein C was also identified in the differential network of our study. Synuclein function is discussed in the earlier part. Overall, we notice enrichment of gene set representing each GO term and apparently a detail investigation is needed to see if newly identified leads in GO term hold any clinical or diagnostic importance. However, enrichment of genes by expanded network and ORA analysis may help us to gain a better insight into the functional role of these proteins.

In general, we notice that the proteome identified in this study is involved in several important immune related responses like apoptosis, phagocytosis, calcium binding, complement activation, negative regulation of blood coagulation and protein kinase activity. Most of these functions are well studied in TB and the current study has tried to populate the associated pathways with the deregulated genes identified either by experimental or *in silico* analysis.

In summary, our experimental findings suggest that the urine proteome is enriched with potential diagnostic markers for different diseases. Additionally, the network based approach provides novel leads by identifying functionally associated molecules and aids biomarker discovery. Previous investigation of some of these molecules in the prospect of being biomarkers of TB augments the efficacy of the network biology method that we adopted here. The putative markers identified in this study either experimentally or by *in silico* analysis may aid investigators to perform targeted validation that may subsequently lead to TB biomarker discovery. We anticipate that our data might contribute to the better understanding of the urine proteome and provide novel leads to TB biomarker discovery. We suggest that proteomics when combined with network biology methods as undertaken in this study could be effectively used to discover biomarkers in other diseases.

## Data Availability

All data produced in the present study are available upon reasonable request to the authors.

## Abbreviations

COPD: Chronic Obstructive Pulmonary Disease
FDR: False Discovery Rate
LC: Lung Cancer
NSCLC: Non-Small Cell Lung Cancer
ORA: Over Representation Analysis
PPI: Protein protein interaction
TB: Tuberculosis
LTBI: Latent TB infection
LOD: Limit of detection.

## Acknowledgement

We acknowledge all the study subjects from Lala Ram Swarup Institute of Tuberculosis and Respiratory Diseases (LRSI) and International Centre for Genetic Engineering and Biotechnology (ICGEB), New Delhi for their kind participation. We are also grateful to Prof. Akhilesh Pandey from the Johns Hopkins University for sharing the data sheet of healthy urine proteome. We thank Dr Saroj Kant Mohapatra from National Institute of Biomedical Genomics, Kalyani for his constructive comments. We thank Sravya Mothe for helping to prepare Figure 3. Contributions of the administrative and support staffs from LRSI are highly acknowledged.

*This work was supported in part by a grant from Department of Biotechnology, Govt. of India (DBT) and infrastructural support of International Centre for Genetic Engineering and Biotechnology, New Delhi.

[S] This article contains Supplementary Tables S1-S7.

## Notes

### Competing Interest Statement

The authors have declared no competing interest.

### Funding Statement

Department of Biotechnology, Government of India

### Author Declarations

Ethics committee and institute review board of International Centre for Genetic Engineering and Biotechnology, New Delhi gave ethical approval for this work. Ethics committee and institute review board of Lala Ram Swarup Institute of Tuberculosis and Respiratory Diseases, New Delhi gave ethical approval for this work.

## References

1. https://pib.gov.in/PressReleasePage.aspx?PRID=1871626

2. Zumla, A., Raviglione, M., Hafner, R., and Fordham von Reyn, C. (2013) Tuberculosis. N. Engl. J. Med. 368, 745–755

3. Shao, C., Li, M., Li, X., Wei, L., Zhu, L., Yang, F., Jia, L., Mu, Y., Wang, J., Guo, Z., Zhang, D., Yin, J., Wang, Z., Sun, W., Zhang, Z., and Gao, Y. (2011) A Tool for Biomarker Discovery in the Urinary Proteome: A Manually Curated Human and Animal Urine Protein Biomarker Database. Mol. Cell. Proteomics 10, M111.010975

4. Banday, K. M., Pasikanti, K. K., Chan, E. C. Y., Singla, R., Rao, K. V. S., Chauhan, V. S., and Nanda, R. K. (2011) Use of Urine Volatile Organic Compounds To Discriminate Tuberculosis Patients from Healthy Subjects. Anal. Chem. 83, 5526–5534

5. Decramer, S., de Peredo, A. G., Breuil, B., Mischak, H., Monsarrat, B., Bascands, J.-L., and Schanstra, J. P. (2008) Urine in Clinical Proteomics. Mol. Cell. Proteomics 7, 1850–1862

6. Shah, M., Variava, E., Holmes, C. B., Coppin, A., Golub, J. E., McCallum, J., Wong, M., Luke, B., Martin, D. J., Chaisson, R. E., Dorman, S. E., and Martinson, N. A. (2009) Diagnostic Accuracy of a Urine Lipoarabinomannan Test for Tuberculosis in Hospitalized Patients in a High HIV Prevalence Setting. J Acquir Immune Defic Syndr 52, 145–151

7. Lawn, S. D., Kerkhoff, A. D., Vogt, M., and Wood, R. (2012) Diagnostic accuracy of a low-cost, urine antigen, point-of-care screening assay for HIV-associated pulmonary tuberculosis before antiretroviral therapy: a descriptive study. Lancet Infect Dis 12, 201–209

8. Afkarian, M., Bhasin, M., Dillon, S. T., Guerrero, M. C., Nelson, R. G., Knowler, W. C., Thadhani, R., and Libermann, T. A. (2010) Optimizing a Proteomics Platform for Urine Biomarker Discovery. Mol. Cell. Proteomics 9, 2195–2204

9. Court, M., Selevsek, N., Matondo, M., Allory, Y., Garin, J., Masselon, C. D., and Domon, B. (2011) Toward a standardized urine proteome analysis methodology. Proteomics 11, 1160–1171

10. Fredolini, C., Meani, F., Alex Reeder, K., Rucker, S., Patanarut, A., Botterell, P., Bishop, B., Longo, C., Espina, V., Petricoin, E., III, Liotta, L., and Luchini, A. (2008) Concentration and preservation of very low abundance biomarkers in urine, such as human growth hormone (hGH), by Cibacron Blue F3G-A loaded hydrogel particles. Nano Research 1, 502–518

11. Ideker, T., and Sharan, R. (2008) Protein networks in disease. Genome Res. 18, 644–652

12. Adachi, J., Kumar, C., Zhang, Y., Olsen, J., and Mann, M. (2006) The human urinary proteome contains more than 1500 proteins, including a large proportion of membrane proteins. Genome Biol 7, R80

13. Li, Q. R., Fan, K. X., Li, R. X., Dai, J., Wu, C. C., Zhao, S. L., Wu, J. R., Shieh, C. H., and Zeng, R. (2010) A comprehensive and non-prefractionation on the protein level approach for the human urinary proteome: touching phosphorylation in urine. Rapid Commun. Mass Spectrom. 24, 823–832

14. Marimuthu, A., O’Meally, R. N., Chaerkady, R., Subbannayya, Y., Nanjappa, V., Kumar, P., Kelkar, D. S., Pinto, S. M., Sharma, R., Renuse, S., Goel, R., Christopher, R., Delanghe, B., Cole, R. N., Harsha, H. C., and Pandey, A. (2011) A Comprehensive Map of the Human Urinary Proteome. J. Proteome Res. 10, 2734–2743

15. Assenov, Y., Ramirez, F., Schelhorn, S.-E., Lengauer, T., and Albrecht, M. (2008) Computing topological parameters of biological networks. Bioinformatics 24, 282–284

16. Shannon, P., Markiel, A., Ozier, O., Baliga, N. S., Wang, J. T., Ramage, D., Amin, N., Schwikowski, B., and Ideker, T. (2003) Cytoscape: A Software Environment for Integrated Models of Biomolecular Interaction Networks. Genome Res. 13, 2498–2504

17. Brohee, S., Faust, K., Lima-Mendez, G., Vanderstocken, G., and van Helden, J. (2008) Network Analysis Tools: from biological networks to clusters and pathways. Nat. Protocols 3, 1616–1629

18. Maere, S., Heymans, K., and Kuiper, M. (2005) BiNGO: a Cytoscape plugin to assess overrepresentation of Gene Ontology categories in Biological Networks. Bioinformatics 21, 3448–3449

19. Merico, D., Isserlin, R., Stueker, O., Emili, A., and Bader, G. D. (2010) Enrichment Map: A Network-Based Method for Gene-Set Enrichment Visualization and Interpretation. PLoS ONE 5, e13984

20. Oesper, L., Merico, D., Isserlin, R., and Bader, G. (2011) WordCloud: a Cytoscape plugin to create a visual semantic summary of networks. Source Code Biol Med 6, 7

21. Chen, E. Y., Xu, H., Gordonov, S., Lim, M. P., Perkins, M. H., and Ma’ayan, A. (2012) Expression2Kinases: mRNA profiling linked to multiple upstream regulatory layers. Bioinformatics 28, 105–111

22. Nepusz, T., Yu, H., and Paccanaro, A. (2012) Detecting overlapping protein complexes in protein-protein interaction networks. Nat Meth 9, 471–472

23. Thomas, C. E., Sexton, W., Benson, K., Sutphen, R., and Koomen, J. (2010) Urine Collection and Processing for Protein Biomarker Discovery and Quantification. Cancer Epidemiol. 19, 953–959

24. Veenstra, T. D. (2007) Global and targeted quantitative proteomics for biomarker discovery. J. Chromatogr., B: Anal. Technol. Biomed. Life Sci. 847, 3–11

25. Liu, Y.-W., Yang, M.-H., Liu, P.-Y., Lee, C.-H., Liao, P.-C., and Tyan, Y.-C. (2008) Proteomic analysis of pericardial effusion: Characteristics of tuberculosis-related proteins. Proteomics Clin Appl 2, 458–466

26. Kentsis, A., Ahmed, S., Kurek, K., Brennan, E., Bradwin, G., Steen, H., and Bachur, R. (2011) Detection and Diagnostic Value of Urine Leucine-Rich alpha 2-Glycoprotein in Children With Suspected Acute Appendicitis. Ann Emerg Med 60, 78–83.e71

27. Huminiecki, L., Gorn, M., Suchting, S., Poulsom, R., and Bicknell, R. (2002) Magic Roundabout Is a New Member of the Roundabout Receptor Family That Is Endothelial Specific and Expressed at Sites of Active Angiogenesis. Genomics 79, 547–552

28. Smith-Berdan, S., Nguyen, A., Hassanein, D., Zimmer, M., Ugarte, F., Ciriza, J. s., Li, D., Garcia-Ojeda, M. E., Hinck, L., and Forsberg, E. C. (2011) Robo4 Cooperates with Cxcr4 to Specify Hematopoietic Stem Cell Localization to Bone Marrow Niches. Cell Stem Cell 8, 72–83

29. Sun, W., Li, F., Wu, S., Wang, X., Zheng, D., Wang, J., and Gao, Y. (2005) Human urine proteome analysis by three separation approaches. Proteomics 5, 4994–5001

30. Edstrom, A. M. L., Malm, J., Frohm, B., Martellini, J. A., Giwercman, A., Morgelin, M., Cole, A. M., and Sorensen, O. E. (2008) The Major Bactericidal Activity of Human Seminal Plasma Is Zinc-Dependent and Derived from Fragmentation of the Semenogelins. J. Immunol. 181, 3413–3421

31. Clemens, D. L., and Horwitz, M. A. (1995) Characterization of the Mycobacterium tuberculosis phagosome and evidence that phagosomal maturation is inhibited. J Exp Med 181, 257–270

32. Lee, B. Y., Jethwaney, D., Schilling, B., Clemens, D. L., Gibson, B. W., and Horwitz, M. A. (2010) The Mycobacterium bovis bacille Calmette-Guerin phagosome proteome. Mol. Cell. Proteomics 9, 32–53

33. Mengaud, J., Ohayon, H., Gounon, P., Mege, R.-M., and Cossart, P. (1996) E-Cadherin Is the Receptor for Internalin, a Surface Protein Required for Entry of L. monocytogenes into Epithelial Cells. Cell 84, 923–932

34. Mohammed, K. A., Nasreen, N., Hardwick, J., Van Horn, R. D., Sanders, K. L., and Antony, V. B. (2003) Mycobacteria induces pleural mesothelial permeability by down-regulating beta-catenin expression. Lung 181, 57–66

35. Jiang, H., Guan, G., Zhang, R., Liu, G., Cheng, J., Hou, X., and Cui, Y. (2009) Identification of urinary soluble E-cadherin as a novel biomarker for diabetic nephropathy. Diabetes Metab Res Rev 25, 232–241

36. Richardson, F., Young, G. D., Sennello, R., Wolf, J., Argast, G. M., Mercado, P., Davies, A., Epstein, D. M., and Wacker, B. (2012) The Evaluation of E-Cadherin and Vimentin as Biomarkers of Clinical Outcomes Among Patients with Non-small Cell Lung Cancer Treated with Erlotinib as Second- or Third-line Therapy. Anticancer Res. 32, 537–552

37. Shariat, S. F., Matsumoto, K., Casella, R., Jian, W., and Lerner, S. P. (2005) Urinary Levels of Soluble E-Cadherin in the Detection of Transitional Cell Carcinoma of the Urinary Bladder. European Urology 48, 69–76

38. Inomata, S. I., Shijubo, N., Kon, S., Maeda, M., Yamada, G., Sato, N., Abe, S., and Uede, T. (2005) Circulating interleukin-18 and osteopontin are useful to evaluate disease activity in patients with tuberculosis. Cytokine 30, 203–211

39. Koguchi, Y., Kawakami, K., Uezu, K., Fukushima, K., Kon, S., Maeda, M., Nakamoto, A., Owan, I., Kuba, M., Kudeken, N., Azuma, M., Yara, S., Shinzato, T., Higa, F., Tateyama, M., Kadota, J.-I., Mukae, H., Kohno, S., Uede, T., and Saito, A. (2003) High Plasma Osteopontin Level and Its Relationship with Interleukin-12-mediated Type 1 T Helper Cell Response in Tuberculosis. Am J Respir Crit Care Med 167, 1355–1359

40. Nau, G. J., Guilfoile, P., Chupp, G. L., Berman, J. S., Kim, S. J., Kornfeld, H., and Young, R. A. (1997) A chemoattractant cytokine associated with granulomas in tuberculosis and silicosis. Proc Natl Acad Sci U S A 94, 6414–6419

41. Bleyer, A. J., Zivna, M., and Kmoch, S. (2011) Uromodulin-associated kidney disease. Nephron Clin Pract 118, c31–36

42. Brown, K. M., Muchmore, A. V., and Rosenstreich, D. L. (1986) Uromodulin, an immunosuppressive protein derived from pregnancy urine, is an inhibitor of interleukin 1. Proc Natl Acad Sci U S A 83, 9119–9123

43. Benetti, E., Caridi, G., Vella, M. D., Rampoldi, L., Ghiggeri, G. M., Artifoni, L., and Murer, L. (2009) Immature Renal Structures Associated With a Novel UMOD Sequence Variant. Am J Kidney Dis 53, 327–331

44. Olakanmi, O., Schlesinger, L. S., Ahmed, A., and Britigan, B. E. (2002) Intraphagosomal Mycobacterium tuberculosis acquires iron from both extracellular transferrin and intracellular iron pools. Impact of interferon-gamma and hemochromatosis. J Biol Chem 277, 49727–49734

45. Hirai, K., Hussey, H. J., Barber, M. D., Price, S. A., and Tisdale, M. J. (1998) Biological Evaluation of a Lipid-mobilizing Factor Isolated from the Urine of Cancer Patients. Cancer Res. 58, 2359–2365

46. Hassan, M. I., Waheed, A., Yadav, S., Singh, T. P., and Ahmad, F. (2008) Zinc alpha 2-glycoprotein: a multidisciplinary protein. Mol Cancer Res 6, 892–906

47. Frank, P. G., and Marcel, Y. L. (2000) Apolipoprotein A-I: structure and function relationships. J. Lipid Res. 41, 853–872

48. Burger, D., and Dayer, J.-M. (2002) High-density lipoprotein-associated apolipoprotein A-I: the missing link between infection and chronic inflammation. Autoimmunity Reviews 1, 111–117

49. Kumar, G. S. S., Venugopal, A. K., Selvan, L. D. N., Marimuthu, A., Keerthikumar, S., Pathare, S., Dikshit, J. B., Tata, P., Hariharan, R., Prasad, T. S. K., Harsha, H. C., Ramachandra, Y. L., Mahadevan, A., Chaerkady, R., Shankar, S. K., and Pandey, A. (2011) Gene Expression Profiling of Tuberculous Meningitis. J Proteomics Bioinform 4, 98–105

50. Neumann, J., Endermann, T., Ehlers, S., and Reiling, N. (2009) Inverse relationship of TLR/NF-kappaB signalling and the Wnt/beta-catenin pathway during inflammation: Deciphering the role of Frizzled1 in M. tuberculosis infection. J Cell Commun Signal 7, A52

51. Maiti, D., Bhattacharyya, A., and Basu, J. (2001) Lipoarabinomannan from Mycobacterium tuberculosis Promotes Macrophage Survival by Phosphorylating Bad through a Phosphatidylinositol 3-Kinase/Akt Pathway. J. Biol. Chem. 276, 329–333

52. Hmama, Z., Sendide, K., Talal, A., Garcia, R., Dobos, K., and Reiner, N. E. (2004) Quantitative analysis of phagolysosome fusion in intact cells: inhibition by mycobacterial lipoarabinomannan and rescue by an 1-alpha-25-dihydroxyvitamin D3-phosphoinositide 3-kinase pathway. J. Cell Sci. 117, 2131–2140

53. Thiel, S. (1992) Mannan-binding protein, a complement activating animal lectin. Immunopharmacology 24, 91–99

54. Selvaraj, P., Narayanan, P. R., and Reetha, A. M. (1999) Association of functional mutant homozygotes of the mannose binding protein gene with susceptibility to pulmonary tuberculosis in India. Tubercle and Lung Disease 79, 221–227

55. Seimon, T. A., Kim, M. J., Blumenthal, A., Koo, J., Ehrt, S., Wainwright, H., Bekker, L. G., Kaplan, G., Nathan, C., Tabas, I., and Russell, D. G. (2010) Induction of ER stress in macrophages of tuberculosis granulomas. PLoS One 5, e12772

56. Zhang, M., Bohlson, S. S., Dy, M., and Tenner, A. J. (2005) Modulated interaction of the ERM protein, moesin, with CD93. Immunology 115, 63–73

57. Good, M. C., Greenstein, A. E., Young, T. A., Ng, H.-L., and Alber, T. (2004) Sensor Domain of the Mycobacterium tuberculosis Receptor Ser/Thr Protein Kinase, PknD, forms a Highly Symmetric beta Propeller. J. Mol. Biol. 339, 459–469

58. Be, N., Bishai, W., and Jain, S. (2012) Role of Mycobacterium tuberculosis pknD in the Pathogenesis of central nervous system tuberculosis. BMC Microbiol 12, 7

59. Steukers, L., Glorieux, S., Vandekerckhove, A. P., Favoreel, H. W., and Nauwynck, H. J. (2012) Diverse microbial interactions with the basement membrane barrier. Trends Microbiol 20, 147–155

60. Monroy, V., Amador, A., Ruiz, B., Espinoza-Cueto, P., Xolalpa, W., Mancilla, R., and Espitia, C. (2000) Binding and activation of human plasminogen by Mycobacterium tuberculosis. Infect Immun 68, 4327–4330

61. Kim, M. J., Wainwright, H. C., Locketz, M., Bekker, L. G., Walther, G. B., Dittrich, C., Visser, A., Wang, W., Hsu, F. F., Wiehart, U., Tsenova, L., Kaplan, G., and Russell, D. G. (2010) Caseation of human tuberculosis granulomas correlates with elevated host lipid metabolism. EMBO Mol Med 2, 258–274

62. Weijer, S., Wieland, C. W., Florquin, S., and van der Poll, T. (2005) A thrombomodulin mutation that impairs activated protein C generation results in uncontrolled lung inflammation during murine tuberculosis. Blood 106, 2761–2768

