## Supplemental Table S1,S2,S3,S4,S5 for "Integrative proteomics and network biology approach to identify potential urine based biomarkers for tuberculosis"

**Supplemental Table S1: Differential expression of proteins observed in LTBI groups**

| Uniprot Accession ID | Protein Description | LTB1-G1 | LTB1-G2 |
|---|---|---|---|
| **P01833** | Polymeric immunoglobulin receptor | 1.20 | 1.53 |
| **Q53H26** | Transferrin variant (Fragment) | 1.74 | 1.32 |
| **Q9UNN8** | Endothelial protein C receptor | 2.50 | - |
| **Q96K68** | cDNA FLJ14473 fis, highly similar to Homo sapiens SNC73 protein | 2.75 | 2.18 |
| **E7EQR8** | Uncharacterized protein GN=YIPF3 | 2.80 | 2.52 |
| **E7ER45** | Uncharacterized protein GN=MGAM | 2.83 | 3.28 |
| **Q6Q3G8** | Lysosomal-associated membrane protein 2 (UNIPROT replaced ID: P13473) | 5.69 | 6.18 |
| **Q96CZ9** | Cadherin 11 | 6.32 | 6.46 |
| **Q4W597** | Osteopontin (UNIPROT replaced ID P10451) | 6.46 | 6.46 |
| P25311 | Zinc-alpha-2-glycoprotein | - | 2.17 |
| **Q6P5S3** | Putative uncharacterized protein PE=2 SV=1 | - | -1.21 |

"- " refers to proteins undetected in the groups.
In case of uncharacterised proteins, the gene name is given by value of "GN"
*Down-regualted are marked in red

**Supplemental Table S2: Differential expression of proteins observed in COPD and Lung Cancer groups**

| Uniprot Accession ID | Description | COPD | Lung cancer |
|---|---|---|---|
| **P01833** | Polymeric immunoglobulin receptor | 1.20 | - |
| **Q9UNN8** | Endothelial protein C receptor | 2.05 | 1.45 |
| **Q96K68** | cDNA FLJ14473 fis, clone MAMMA1001080, highly similar to Homo sapiens SNC73 protein | 2.26 | - |
| **P25311** | Zinc-alpha-2-glycoprotein | 2.51 | - |
| **P02760** | Protein AMBP | 2.56 | - |
| **B3KRN9** | cDNA FLJ34642 highly similar to Uromodulin (UNIPROT replaced ID: P07911-Uromodulin) | 2.71 | -1.69 |
| **E7EQR8** | Uncharacterized protein GN=YIPF3 | 2.80 | - |
| **Q53H26** | Transferrin variant (Fragment) | 2.92 | - |
| **Q96CZ9** | Cadherin 11 | 6.40 | - |
| **Q4W597** | Putative uncharacterized protein SPP1 (UNIPROT replaced ID P10451-osteopontin) | 6.44 | 5.86 |
| **Q6Q3G8** | Lysosomal-associated membrane protein 2 (UNIPROT replaced ID: P13473 -Lysosome-associated membrane glycoprotein 2) | 6.46 | 5.87 |
| **E7ER45** | Uncharacterized protein GN=MGAM | 1.36 | -3.27 |
| **B2R888** | Monocyte differentiation antigen CD14 | - | -5.13 |
| **B7Z545** | cDNA FLJ60068, highly similar to Inter-alpha-trypsin inhibitor heavy chain H4 (UNIPROT replaced ID:Q14624-Inter-alpha-trypsin inhibitor heavy chain H4) | - | -5.83 |
| **O00187** | Mannan-binding lectin serine protease 2 | - | -4.56 |
| **P01008** | Antithrombin-III | - | -1.57 |
| **P01034** | Cystatin-C | - | -6.55 |
| **P06870** | Kallikrein-1 | - | -2.67 |
| **P16070** | CD44 antigen | - | -3.11 |
| **P63261** | Actin | - | -1.94 |
| **Q7Z3B1** | Neuronal growth regulator 1 | - | -6.50 |
| **Q9UL83** | Myosin-reactive immunoglobulin light chain variable region (Fragment) | - | -6.48 |

*Down-regulated proteins are marked in red

### Supplemental Table S3: Differential expression of proteins observed in two TB groups

| Uniprot Accession | Name | Fold change (TB Group 1) | Fold change (TB Group 2) |
|---|---|---|---|
| **Q14204** | Cytoplasmic dynein 1 heavy chain 1 | -6.07 | -3.77 |
| **B2R582** | cDNA, FLJ92374, highly similar to Homo sapiens C-type lectin domain family 3, member B (CLEC3B), mRNA PE=2 SV=1 | -4.41 | -3.95 |
| **A2NUT2** | Lambda-chain (AA -20 to 215) | -4.31 | -3.08 |
| **B2RDA1** | Secreted phosphoprotein 1 (Osteopontin) | -3.63 | -2.68 |
| **B7ZL69** | CREB3L3 protein | -3.61 | -2.96 |
| **P02671** | Fibrinogen alpha chain | -2.70 | -4.08 |
| **P05154** | Plasma serine protease inhibitor | -2.38 | -2.26 |
| **Q14508** | WAP four-disulfide core domain protein 2 | -2.38 | -2.55 |
| **Q6FGN3** | Syndecan | -2.30 | -2.05 |
| **P98160** | Basement membrane-specific heparan sulfate proteoglycan core protein | -1.81 | -2.48 |
| **Q14515** | SPARC-like protein 1 | -1.81 | -2.14 |
| **E3SFM9** | Neuregulin 1 type IV beta 1a | -1.55 | -1.58 |
| **P02452** | Collagen alpha-1(I) chain | -1.50 | - |
| **Q6LBL5** | GM2 activator protein | -1.37 | -2.15 |
| **P01625** | Ig kappa chain V-IV region Len | -1.36 | - |
| **Q5U050** | Ciliary neurotrophic factor receptor | -1.26 | -1.79 |
| **Q76LA1** | CSTB protein | -1.08 | -1.42 |
| **B2R5M3** | cDNA, FLJ92530, highly similar to Homo sapiens chromogranin B (secretogranin 1) (CHGB), mRNA PE=2 SV=1 | -1.04 | -1.42 |
| **P63261** | Actin | 1.10 | 1.20 |
| **B7Z8Q2** | cDNA FLJ55606, highly similar to Alpha-2-HS-glycoprotein PE=2 SV=1 | 1.13 | - |
| **P01617** | Ig kappa chain V-II region | 1.13 | - |
| **E7EW61** | Uncharacterized protein GN=TTR | 1.13 | - |
| **P01009** | Alpha-1-antitrypsin | 1.18 | 1.40 |
| **E7EUP8** | Uncharacterized protein GN=PGA3 | 1.24 | 2.10 |
| **D9YZU5** | Hemoglobin, beta | 1.33 | - |
| **P02647** | Apolipoprotein A-I | 1.40 | 1.26 |
| **E7EVV1** | Uncharacterized protein GN=HP | 1.40 | - |
| **O94822** | E3 ubiquitin-protein ligase listerin GN=LTN1 | 1.46 | 1.58 |
| **Q8N114** | Protein shisa-5 | 1.61 | 1.74 |
| **P02750** | Leucine-rich alpha-2-glycoprotein | 1.62 | 1.33 |
| **A8MWK3** | Uncharacterized protein GN=CDH2 | 1.62 | 1.02 |
| **B2R9F2** | cDNA, FLJ94361, highly similar to Homo sapiens serine (or cysteine) proteinase inhibitor, clade A(alpha-1 antiproteinase, antitrypsin), member 6 (SERPINA6), mRNA PE=2 SV=1 | 1.65 | - |
| **Q60FE6** | Filamin A | 1.65 | 2.01 |
| **Q9UII7** | E-cadherin | 1.69 | 1.55 |
| **A8K6V6** | cDNA FLJ75883, highly similar to Homo sapiens glucosamine (N-acetyl)-6-sulfatase (Sanfilippo disease IIID) (GNS), | 1.70 | - |
| **P02790** | Hemopexin | 1.77 | - |
| **P01833** | Polymeric immunoglobulin receptor | 1.82 | 1.63 |
| **E7ER45** | Uncharacterized protein GN=MGAM | 1.86 | 1.25 |
| **B7Z6N2** | cDNA FLJ56154, highly similar to Gelsolin | 1.91 | 1.14 |
| **Q96K68** | cDNA FLJ14473 fis, clone MAMMA1001080, highly similar to Homo sapiens SNC73 protein (SNC73) mRNA | 1.95 | - |
| **P04217** | Alpha-1B-glycoprotein | 1.99 | 1.21 |
| **B3KS79** | cDNA FLJ35730 fis, clone TESTI2003131, highly similar to ALPHA-1-ANTICHYMOTRYPSIN | 2.05 | 1.95 |
| **O00187** | Mannan-binding lectin serine protease 2 | 2.06 | 2.21 |
| **P07998** | Ribonuclease pancreatic | 2.06 | 2.14 |
| **P01042-2** | Isoform LMW of Kininogen-1 | 2.09 | 2.82 |
| **P10909-2** | Isoform 2 of Clusterin | 2.10 | 2.51 |
| **P04279** | Semenogelin-1 | 2.15 | 1.41 |
| **Q6FHM9** | CD59 antigen, complement regulatory protein | 2.21 | 1.89 |
| **Q6LDS3** | APS protein (Fragment) | 2.22 | 1.38 |
| **B7Z545** | cDNA FLJ60068, highly similar to Inter-alpha-trypsin inhibitor heavy chain H4 PE=2 SV=1 | 2.22 | 1.32 |
| **D3DPK5** | SH3 domain binding glutamic acid-rich protein like 3, isoform CRA_a (Fragment) | 2.23 | 1.28 |
| **Q6IRW3** | Semenogelin II | 2.25 | 2.56 |
| **Q9UL85** | Myosin-reactive immunoglobulin kappa chain variable region (Fragment) | 2.26 | 2.31 |

| | | | |
|---|---|---|---|
| **B4DVE1** | cDNA FLJ53478, highly similar to Galectin-3-binding protein | 2.27 | 1.44 |
| **C9JF17** | Uncharacterized protein GN=APOD | 2.29 | - |
| **P06870** | Kallikrein-1 | 2.31 | 1.87 |
| **E7EVD2** | Uncharacterized protein GN=EGF | 2.35 | 1.93 |
| **P02760** | Protein AMBP | 2.39 | 1.94 |
| **P25311** | Zinc-alpha-2-glycoprotein | 2.51 | 2.07 |
| **Q53H26** | Transferrin variant (Fragment) | 2.60 | 1.49 |
| **D6RBV2** | Uncharacterized protein | 2.78 | 3.18 |
| **P02768** | Serum albumin | 2.84 | 2.75 |
| **P01871** | Ig mu chain C region | 2.94 | 2.95 |
| **P04433** | Ig kappa chain V-III region VG (Fragment) | 2.99 | 3.20 |
| **D3YTG1** | Inducible T-cell co-stimulator ligand | 3.04 | 3.30 |
| **B3KRN9** | cDNA FLJ34642 fis, clone KIDNE2016918, highly similar to UROMODULIN PE=2 SV=1 | 3.06 | 2.76 |
| **E7EQR8** | Uncharacterized protein GN=YIPF3 | 3.10 | 2.92 |
| **Q6PJF2** | IGK@ protein GN=IGK@ | 3.18 | 2.37 |
| **P08637** | Low affinity immunoglobulin gamma Fc region receptor III-A GN=FCGR3A | 3.19 | 3.24 |
| **A2KBC7** | Anti-IFN-G scFv (Fragment) PE=2 SV=1 | 3.24 | 3.53 |
| **P11279** | Lysosome-associated membrane glycoprotein 1 GN=LAMP1 PE=1 SV=3 | 3.41 | 3.91 |
| **P00734** | Prothrombin | 3.41 | 2.56 |
| **B2R5L2** | cDNA, FLJ92516, highly similar to Homo sapiens orosomucoid 2 (ORM2), mRNA PE=2 SV=1 | 3.44 | 1.87 |
| **A8K460** | cDNA FLJ78736, highly similar to Homo sapiens phospholipase A2, group VI (cytosolic, calcium-independent) (PLA2G6), transcript variant 2, mRNA PE=2 SV=1 | 3.60 | 5.89 |
| **Q6N093** | Putative uncharacterized protein DKFZp686I04196 (Fragment) GN=DKFZp686I04196 | 3.60 | 4.28 |
| **Q5T539** | Orosomucoid 1 | 4.33 | 2.83 |
| **A1A508** | PRSS3 protein | 5.09 | 5.43 |
| **P08294** | Extracellular superoxide dismutase [Cu-Zn] | 5.26 | 4.70 |
| **Q96CZ9** | Cadherin 11, type 2, OB-cadherin (Osteoblast) | 6.32 | 6.34 |
| **Q4W597** | Putative uncharacterized protein SPP1 | 6.43 | 6.23 |
| **Q6Q3G8** | Lysosomal-associated membrane protein 2 | 6.46 | 6.46 |
| **Q9HD89** | Resistin | 6.63 | 6.63 |
| **A6XND9** | Beta-2-microglobulin | - | -1.30 |
| **P16070** | CD44 antigen GN=CD44 | - | -1.28 |
| **P04264** | Keratin, type II cytoskeletal 1 | - | 1.36 |
| **P10153** | Non-secretory ribonuclease | - | -1.08 |
| **P41222** | Prostaglandin-H2 D-isomerase | - | -1.14 |
| **Q5EBM2** | Putative uncharacterized protein | - | 2.62 |
| **Q8WZ75** | Roundabout homolog 4 | - | -4.15 |
| **P05543** | Thyroxine-binding globulin | - | 1.05 |
| **P30530** | Tyrosine-protein kinase receptor UFO | - | -1.99 |
| **D6RAK8** | Uncharacterized protein GN=GC | - | -1.49 |

"- " refers to proteins undetected in the groups. In case of uncharacterised proteins, the gene name is given by value of "GN"

### Supplemental Table S4: Over-representation of gene onotological terms observed in the up-regulated gene set identified in TB groups

| GO-ID | p-value | corr p-value | x | n | X | N | Description | Genes in test set |
|---|---|---|---|---|---|---|---|---|
| **5576** | 3.4369E-24 | 4.7602E-21 | 38 | 2026 | 54 | 17786 | extracellular region | TF\|MASP2\|UMOD\|IGHM\|AHSG\|AZGP1\|LGALS3BP\|APOA1\|APOD\|ALB\|LRG1\|SERPINA6\|GSN\|PRSS3\|APS\|ITIH4\|SERPINA3\|SERPINA1\|EGF\|FCGR3A\|SPP1\|KNG1\|RNASE1\|SEMG1\|SEMG2\|PIGR\|SOD3\|FLNA\|A1BG\|AMBP\|DKFZP686I04196\|ORM1\|RETN\|HPX\|CD59\|F2\|IGKV3-20\|ORM2 |
| **5615** | 1.3072E-15 | 9.0521E-13 | 21 | 747 | 54 | 17786 | extracellular space | KNG1\|TF\|SEMG1\|SEMG2\|SOD3\|AHSG\|ORM1\|RETN\|LGALS3BP\|APOA1\|APOD\|HPX\|ALB\|GSN\|SERPINA6\|PRSS3\|F2\|SERPINA1\|EGF\|ORM2\|SPP1 |
| **44421** | 3.0009E-13 | 1.3854E-10 | 21 | 985 | 54 | 17786 | extracellular region part | KNG1\|TF\|SEMG1\|SEMG2\|SOD3\|AHSG\|ORM1\|RETN\|LGALS3BP\|APOA1\|APOD\|HPX\|ALB\|GSN\|SERPINA6\|PRSS3\|F2\|SERPINA1\|EGF\|ORM2\|SPP1 |
| **2526** | 5.7485E-12 | 1.9904E-09 | 9 | 89 | 54 | 17786 | acute inflammatory response | ORM1\|TF\|MASP2\|F2\|ITIH4\|SERPINA3\|SERPINA1\|ORM2\|AHSG |
| **6953** | 1.8642E-11 | 5.1639E-09 | 7 | 38 | 54 | 17786 | acute-phase response | ORM1\|F2\|ITIH4\|SERPINA3\|SERPINA1\|ORM2\|AHSG |
| **6954** | 2.1955E-09 | 5.0679E-07 | 11 | 315 | 54 | 17786 | inflammatory response | KNG1\|ORM1\|TF\|MASP2\|F2\|ITIH4\|SERPINA3\|SERPINA1\|ORM2\|SPP1\|AHSG |
| **9611** | 5.8461E-09 | 1.1567E-06 | 13 | 541 | 54 | 17786 | response to wounding | KNG1\|ORM1\|TF\|GSN\|MASP2\|CD59\|F2\|ITIH4\|SERPINA3\|SERPINA1\|ORM2\|SPP1\|AHSG |
| **6952** | 2.9458E-08 | 5.0998E-06 | 13 | 620 | 54 | 17786 | defense response | KNG1\|ORM1\|TF\|LGALS3BP\|MASP2\|F2\|ITIH4\|SERPINA3\|UMOD\|SERPINA1\|ORM2\|SPP1\|AHSG |
| **30141** | 8.4875E-08 | 0.000013061 | 8 | 186 | 54 | 17786 | stored secretory granule | TF\|LAMP2\|APOA1\|SEMG1\|ALB\|SEMG2\|SERPINA1\|EGF |
| **4866** | 2.7882E-07 | 0.000036776 | 7 | 146 | 54 | 17786 | endopeptidase inhibitor activity | AMBP\|KNG1\|SERPINA6\|ITIH4\|SERPINA3\|SERPINA1\|AHSG |
| **61135** | 2.9208E-07 | 0.000036776 | 7 | 147 | 54 | 17786 | endopeptidase regulator activity | AMBP\|KNG1\|SERPINA6\|ITIH4\|SERPINA3\|SERPINA1\|AHSG |
| **30414** | 4.1864E-07 | 0.000048318 | 7 | 155 | 54 | 17786 | peptidase inhibitor activity | AMBP\|KNG1\|SERPINA6\|ITIH4\|SERPINA3\|SERPINA1\|AHSG |
| **61134** | 1.0644E-06 | 0.0001134 | 7 | 178 | 54 | 17786 | peptidase regulator activity | AMBP\|KNG1\|SERPINA6\|ITIH4\|SERPINA3\|SERPINA1\|AHSG |
| **6950** | 2.5087E-06 | 0.00024818 | 18 | 1773 | 54 | 17786 | response to stress | KNG1\|TF\|MASP2\|CDH1\|UMOD\|SOD3\|AHSG\|ORM1\|LGALS3BP\|ALB\|GSN\|CD59\|F2\|SERPINA3\|ITIH4\|SERPINA1\|ORM2\|SPP1 |
| **48583** | 3.3737E-06 | 0.0003115 | 10 | 524 | 54 | 17786 | regulation of response to stimulus | AMBP\|KNG1\|TF\|APOA1\|HPX\|MASP2\|F2\|APS\|SPP1\|AHSG |
| **31988** | 4.0605E-06 | 0.00035149 | 11 | 665 | 54 | 17786 | membrane-bounded vesicle | TF\|LAMP2\|APOA1\|SEMG1\|ALB\|SEMG2\|YIPF3\|SERPINA1\|PIGR\|EGF\|SPP1 |
| **50896** | 4.6996E-06 | 0.00038288 | 26 | 3632 | 54 | 17786 | response to stimulus | TF\|MASP2\|CDH1\|UMOD\|IGHM\|AHSG\|AZGP1\|LGALS3BP\|GSN\|ALB\|APS\|ITIH4\|SERPINA3\|IGHA1\|SERPINA1\|FCGR3A\|SPP1\|KNG1\|SOD3\|RETN\|ORM1\|DKFZP686I04196\|CD59\|F2\|IGKV3-20\|ORM2 |
| **31982** | 7.9865E-06 | 0.00059883 | 11 | 714 | 54 | 17786 | vesicle | TF\|LAMP2\|APOA1\|SEMG1\|ALB\|SEMG2\|YIPF3\|SERPINA1\|PIGR\|EGF\|SPP1 |
| **65008** | 0.000008215 | 0.00059883 | 16 | 1541 | 54 | 17786 | regulation of biological quality | KNG1\|TF\|FLNA\|APOA1\|HPX\|GSN\|SERPINA6\|ALB\|CD59\|APS\|F2\|SERPINA3\|SERPINA1\|SH3BGRL3\|HBB\|SPP1 |
| **4867** | 0.000009058 | 0.00062727 | 5 | 93 | 54 | 17786 | serine-type endopeptidase inhibitor activity | AMBP\|SERPINA6\|ITIH4\|SERPINA3\|SERPINA1 |
| **4857** | 0.00002044 | 0.0013414 | 7 | 279 | 54 | 17786 | enzyme inhibitor activity | AMBP\|KNG1\|SERPINA6\|ITIH4\|SERPINA3\|SERPINA1\|AHSG |

| | | | | | | | | |
|---|---|---|---|---|---|---|---|---|
| **16023** | 0.000021308 | 0.0013414 | 10 | 647 | 54 | 17786 | cytoplasmic membrane-bounded vesicle | TF\|LAMP2\|APOA1\|SEMG1\|ALB\|SEMG2\|YIPF3\|SERPINA1\|PIGR\|EGF |
| **3823** | 0.000030513 | 0.0018374 | 4 | 59 | 54 | 17786 | antigen binding | DKFZP686I04196\|IGHA1\|IGKV3-20\|IGHM |
| **31410** | 0.000034687 | 0.0020017 | 10 | 685 | 54 | 17786 | cytoplasmic vesicle | TF\|LAMP2\|APOA1\|SEMG1\|ALB\|SEMG2\|YIPF3\|SERPINA1\|PIGR\|EGF |
| **80134** | 0.00004804 | 0.0026614 | 7 | 319 | 54 | 17786 | regulation of response to stress | AMBP\|KNG1\|TF\|HPX\|F2\|SPP1\|AHSG |
| **50878** | 0.000079616 | 0.0042411 | 5 | 146 | 54 | 17786 | regulation of body fluid levels | KNG1\|TF\|CD59\|F2\|SERPINA1 |
| **42153** | 0.000089949 | 0.004614 | 2 | 5 | 54 | 17786 | RPTP-like protein binding | CDH1\|CDH2 |
| **5509** | 0.00010091 | 0.0049915 | 9 | 626 | 54 | 17786 | calcium ion binding | GSN\|MASP2\|PRSS3\|F2\|UMOD\|CDH1\|CDH2\|EGF\|CDH11 |
| **4252** | 0.00010561 | 0.0050437 | 5 | 155 | 54 | 17786 | serine-type endopeptidase activity | MASP2\|PRSS3\|F2\|APS\|KLK1 |
| **31093** | 0.00016164 | 0.0074623 | 3 | 35 | 54 | 17786 | platelet alpha granule lumen | ALB\|SERPINA1\|EGF |
| **32880** | 0.00017202 | 0.0076155 | 5 | 172 | 54 | 17786 | regulation of protein localization | APOA1\|CDH1\|CDH2\|EGF\|FLNA |
| **60205** | 0.00017595 | 0.0076155 | 3 | 36 | 54 | 17786 | cytoplasmic membrane-bounded vesicle lumen | ALB\|SERPINA1\|EGF |
| **16342** | 0.00018816 | 0.0078969 | 2 | 7 | 54 | 17786 | catenin complex | CDH1\|CDH2 |
| **8236** | 0.00019659 | 0.0080081 | 5 | 177 | 54 | 17786 | serine-type peptidase activity | MASP2\|PRSS3\|F2\|APS\|KLK1 |
| **31983** | 0.000207 | 0.0081778 | 3 | 38 | 54 | 17786 | vesicle lumen | ALB\|SERPINA1\|EGF |
| **17171** | 0.00021256 | 0.0081778 | 5 | 180 | 54 | 17786 | serine hydrolase activity | MASP2\|PRSS3\|F2\|APS\|KLK1 |
| **30193** | 0.00022377 | 0.0083303 | 3 | 39 | 54 | 17786 | regulation of blood coagulation | KNG1\|TF\|F2 |
| **30234** | 0.00022856 | 0.0083303 | 10 | 860 | 54 | 17786 | enzyme regulator activity | AMBP\|KNG1\|APOA1\|SERPINA6\|ITIH4\|SERPINA3\|CDH1\|SERPINA1\|EGF\|AHSG |
| **8211** | 0.00025039 | 0.0086697 | 2 | 8 | 54 | 17786 | glucocorticoid metabolic process | APOA1\|SERPINA6 |
| **31233** | 0.00025039 | 0.0086697 | 2 | 8 | 54 | 17786 | intrinsic to external side of plasma membrane | TF\|CD59 |
| **50817** | 0.00026855 | 0.0088274 | 4 | 103 | 54 | 17786 | coagulation | TF\|CD59\|F2\|SERPINA1 |
| **7596** | 0.00026855 | 0.0088274 | 4 | 103 | 54 | 17786 | blood coagulation | TF\|CD59\|F2\|SERPINA1 |
| **32101** | 0.00027975 | 0.0088274 | 5 | 191 | 54 | 17786 | regulation of response to external stimulus | KNG1\|TF\|F2\|SPP1\|AHSG |
| **2682** | 0.00028044 | 0.0088274 | 7 | 424 | 54 | 17786 | regulation of immune system process | AMBP\|ORM1\|APOA1\|HPX\|MASP2\|APS\|ORM2 |
| **61041** | 0.00029959 | 0.0088284 | 3 | 43 | 54 | 17786 | regulation of wound healing | KNG1\|TF\|F2 |
| **50818** | 0.00029959 | 0.0088284 | 3 | 43 | 54 | 17786 | regulation of coagulation | KNG1\|TF\|F2 |
| **90316** | 0.00029959 | 0.0088284 | 3 | 43 | 54 | 17786 | positive regulation of intracellular protein transport | CDH1\|EGF\|FLNA |
| **7599** | 0.00033341 | 0.0095519 | 4 | 109 | 54 | 17786 | hemostasis | TF\|CD59\|F2\|SERPINA1 |
| **42060** | 0.00033794 | 0.0095519 | 5 | 199 | 54 | 17786 | wound healing | TF\|GSN\|CD59\|F2\|SERPINA1 |
| **32388** | 0.00036615 | 0.010142 | 3 | 46 | 54 | 17786 | positive regulation of intracellular transport | CDH1\|EGF\|FLNA |
| **45177** | 0.00040488 | 0.010995 | 5 | 207 | 54 | 17786 | apical part of cell | TF\|MGAM\|UMOD\|CDH1\|SPP1 |
| **45295** | 0.00048897 | 0.012986 | 2 | 11 | 54 | 17786 | gamma-catenin binding | CDH1\|CDH2 |
| **31091** | 0.00049695 | 0.012986 | 3 | 51 | 54 | 17786 | platelet alpha granule | ALB\|SERPINA1\|EGF |
| **45444** | 0.00065444 | 0.016785 | 3 | 56 | 54 | 17786 | fat cell differentiation | RETN\|LRG1\|APS |
| **50776** | 0.00072032 | 0.018139 | 5 | 235 | 54 | 17786 | regulation of immune response | AMBP\|APOA1\|HPX\|MASP2\|APS |

| | | | | | | | | |
|---|---|---|---|---|---|---|---|---|
| **48519** | 0.00074393 | 0.018399 | 15 | 20 20 | 54 | 17786 | negative regulation of biological process | KNG1\|TF\|CDH1\|UMOD\|FLNA\|AHSG\|AMBP\|AZGP1\|APOA1\|ALB\|GSN\|APS\|F2\|EGF\|SPP1 |
| **10035** | 0.00076265 | 0.018531 | 5 | 23 8 | 54 | 17786 | response to inorganic substance | TF\|GSN\|CDH1\|SERPINA1\|SOD3 |
| **30194** | 0.00080431 | 0.018881 | 2 | 14 | 54 | 17786 | positive regulation of blood coagulation | TF\|F2 |
| **34394** | 0.00080431 | 0.018881 | 2 | 14 | 54 | 17786 | protein localization at cell surface | EGF\|FLNA |
| **51223** | 0.00097679 | 0.022548 | 4 | 14 5 | 54 | 17786 | regulation of protein transport | APOA1\|CDH1\|EGF\|FLNA |
| **42306** | 0.0010116 | 0.022967 | 3 | 65 | 54 | 17786 | regulation of protein import into nucleus | CDH1\|EGF\|FLNA |
| **70201** | 0.001192 | 0.025078 | 4 | 15 3 | 54 | 17786 | regulation of establishment of protein localization | APOA1\|CDH1\|EGF\|FLNA |
| **10038** | 0.001192 | 0.025078 | 4 | 15 3 | 54 | 17786 | response to metal ion | GSN\|CDH1\|SERPINA1\|SOD3 |
| **19865** | 0.001195 | 0.025078 | 2 | 17 | 54 | 17786 | immunoglobulin binding | AMBP\|FCGR3A |
| **50820** | 0.001195 | 0.025078 | 2 | 17 | 54 | 17786 | positive regulation of coagulation | TF\|F2 |
| **18298** | 0.001195 | 0.025078 | 2 | 17 | 54 | 17786 | protein-chromophore linkage | AMBP\|IGHA1 |
| **51239** | 0.0012359 | 0.025549 | 10 | 10 67 | 54 | 17786 | regulation of multicellular organismal process | KNG1\|TF\|APOA1\|F2\|APS\|CDH1\|CDH2\|EGF\|SPP1\|AHSG |
| **51050** | 0.0013387 | 0.026933 | 5 | 27 0 | 54 | 17786 | positive regulation of transport | F2\|CDH1\|EGF\|FLNA\|AHSG |
| **42993** | 0.0013418 | 0.026933 | 2 | 18 | 54 | 17786 | positive regulation of transcription factor import into nucleus | CDH1\|FLNA |
| **32879** | 0.001476 | 0.028979 | 8 | 72 7 | 54 | 17786 | regulation of localization | TF\|APOA1\|F2\|CDH1\|CDH2\|EGF\|FLNA\|AHSG |
| **8289** | 0.0014856 | 0.028979 | 6 | 41 1 | 54 | 17786 | lipid binding | TF\|AZGP1\|APOA1\|APOD\|ALB\|SERPINA6 |
| **33157** | 0.0017773 | 0.034189 | 3 | 79 | 54 | 17786 | regulation of intracellular protein transport | CDH1\|EGF\|FLNA |
| **51605** | 0.0019094 | 0.035781 | 3 | 81 | 54 | 17786 | protein maturation by peptide bond cleavage | TF\|MASP2\|PRSS3 |
| **51241** | 0.0019118 | 0.035781 | 4 | 17 4 | 54 | 17786 | negative regulation of multicellular organismal process | KNG1\|APOA1\|F2\|AHSG |
| **46822** | 0.0019777 | 0.036522 | 3 | 82 | 54 | 17786 | regulation of nucleocytoplasmic transport | CDH1\|EGF\|FLNA |
| **22603** | 0.0021559 | 0.037545 | 5 | 30 1 | 54 | 17786 | regulation of anatomical structure morphogenesis | TF\|APS\|CDH1\|CDH2\|SPP1 |
| **9966** | 0.0021713 | 0.037545 | 8 | 77 3 | 54 | 17786 | regulation of signal transduction | AMBP\|TF\|HPX\|SHISA5\|APS\|CDH2\|EGF\|FLNA |
| **42592** | 0.0021889 | 0.037545 | 8 | 77 4 | 54 | 17786 | homeostatic process | KNG1\|TF\|APOA1\|HPX\|F2\|APS\|SERPINA3\|SH3BGRL3 |
| **51222** | 0.0021916 | 0.037545 | 3 | 85 | 54 | 17786 | positive regulation of protein transport | CDH1\|EGF\|FLNA |
| **42307** | 0.0021973 | 0.037545 | 2 | 23 | 54 | 17786 | positive regulation of protein import into nucleus | CDH1\|FLNA |
| **23051** | 0.0022603 | 0.037545 | 8 | 77 8 | 54 | 17786 | regulation of signaling process | AMBP\|TF\|HPX\|SHISA5\|APS\|CDH2\|EGF\|FLNA |
| **30195** | 0.0023925 | 0.037545 | 2 | 24 | 54 | 17786 | negative regulation of blood coagulation | KNG1\|F2 |
| **50793** | 0.0025052 | 0.037545 | 8 | 79 1 | 54 | 17786 | regulation of developmental process | TF\|F2\|APS\|CDH1\|CDH2\|EGF\|SPP1\|AHSG |
| **6955** | 0.0025591 | 0.037545 | 7 | 61 9 | 54 | 17786 | immune response | DKFZP686I04196\|AZGP1\|MASP2\|IGHA1\|IGKV3-20\|FCGR3A\|IGHM |
| **19898** | 0.0025791 | 0.037545 | 3 | 90 | 54 | 17786 | extrinsic to membrane | UMOD\|CDH1\|CDH2 |
| **43542** | 0.0025954 | 0.037545 | 2 | 25 | 54 | 17786 | endothelial cell migration | APOA1\|PRSS3 |
| **50873** | 0.0025954 | 0.037545 | 2 | 25 | 54 | 17786 | brown fat cell differentiation | LRG1\|APS |

| | | | | | | | | |
|---|---|---|---|---|---|---|---|---|
| **43281** | 0.0027452 | 0.037545 | 3 | 92 | 54 | 17786 | regulation of caspase activity | TF\|F2\|CDH1 |
| **50819** | 0.0028063 | 0.037545 | 2 | 26 | 54 | 17786 | negative regulation of coagulation | KNG1\|F2 |
| **10627** | 0.0029241 | 0.037545 | 5 | 32 3 | 54 | 17786 | regulation of intracellular protein kinase cascade | AMBP\|TF\|HPX\|SHISA5\|FLNA |
| **35466** | 0.0029887 | 0.037545 | 9 | 10 03 | 54 | 17786 | regulation of signaling pathway | AMBP\|TF\|HPX\|SHISA5\|APS\|CDH2\|EGF\|FLNA\|AHSG |
| **34987** | 0.0030361 | 0.037545 | 1 | 1 | 54 | 17786 | immunoglobulin receptor binding | FLNA |
| **34989** | 0.0030361 | 0.037545 | 1 | 1 | 54 | 17786 | GTP-Ral binding | FLNA |
| **34988** | 0.0030361 | 0.037545 | 1 | 1 | 54 | 17786 | Fc-gamma receptor I complex binding | FLNA |
| **2543** | 0.0030361 | 0.037545 | 1 | 1 | 54 | 17786 | activation of blood coagulation via clotting cascade | TF |
| **8449** | 0.0030361 | 0.037545 | 1 | 1 | 54 | 17786 | N-acetylglucosamine-6-sulfatase activity | GNS |
| **50713** | 0.0030361 | 0.037545 | 1 | 1 | 54 | 17786 | negative regulation of interleukin-1 beta secretion | APOA1 |
| **46687** | 0.0030361 | 0.037545 | 1 | 1 | 54 | 17786 | response to chromate | SERPINA1 |
| **48685** | 0.0030361 | 0.037545 | 1 | 1 | 54 | 17786 | negative regulation of collateral sprouting of intact axon in response to injury | SPP1 |
| **48683** | 0.0030361 | 0.037545 | 1 | 1 | 54 | 17786 | regulation of collateral sprouting of intact axon in response to injury | SPP1 |
| **60330** | 0.0030361 | 0.037545 | 1 | 1 | 54 | 17786 | regulation of response to interferon-gamma | HPX |
| **60335** | 0.0030361 | 0.037545 | 1 | 1 | 54 | 17786 | positive regulation of interferon-gamma-mediated signaling pathway | HPX |
| **60332** | 0.0030361 | 0.037545 | 1 | 1 | 54 | 17786 | positive regulation of response to interferon-gamma | HPX |
| **34191** | 0.0030361 | 0.037545 | 1 | 1 | 54 | 17786 | apolipoprotein A-I receptor binding | APOA1 |
| **46904** | 0.0030361 | 0.037545 | 1 | 1 | 54 | 17786 | calcium oxalate binding | AMBP |
| **33986** | 0.0030361 | 0.037545 | 1 | 1 | 54 | 17786 | response to methanol | SERPINA1 |
| **15886** | 0.0030361 | 0.037545 | 1 | 1 | 54 | 17786 | heme transport | HPX |
| **34014** | 0.0030361 | 0.037545 | 1 | 1 | 54 | 17786 | response to triglyceride | SERPINA1 |
| **5982** | 0.0030361 | 0.037545 | 1 | 1 | 54 | 17786 | starch metabolic process | MGAM |
| **5983** | 0.0030361 | 0.037545 | 1 | 1 | 54 | 17786 | starch catabolic process | MGAM |
| **51764** | 0.0030361 | 0.037545 | 1 | 1 | 54 | 17786 | actin crosslink formation | FLNA |
| **1922** | 0.0030361 | 0.037545 | 1 | 1 | 54 | 17786 | B-1 B cell homeostasis | APS |
| **32386** | 0.0031889 | 0.038487 | 3 | 97 | 54 | 17786 | regulation of intracellular transport | CDH1\|EGF\|FLNA |
| **35239** | 0.0032204 | 0.038487 | 4 | 20 1 | 54 | 17786 | tube morphogenesis | KNG1\|CDH1\|EGF\|HBB |
| **272** | 0.0032513 | 0.038487 | 2 | 28 | 54 | 17786 | polysaccharide catabolic process | GNS\|MGAM |
| **2020** | 0.0032513 | 0.038487 | 2 | 28 | 54 | 17786 | protease binding | TF\|SERPINA1 |
| **42168** | 0.0032513 | 0.038487 | 2 | 28 | 54 | 17786 | heme metabolic process | AMBP\|HPX |
| **52548** | 0.0032826 | 0.038529 | 3 | 98 | 54 | 17786 | regulation of endopeptidase activity | TF\|F2\|CDH1 |
| **8150** | 0.0034932 | 0.040657 | 51 | 14 30 2 | 54 | 17786 | biological_process | TF\|MASP2\|UMOD\|CDH1\|LMAN2\|CDH2\|IGHM\|AHSG\|ACTG1\|AZGP1\|LGALS3BP\|APOA1\|APOD\|ALB\|LRG1\|SERPINA6\|GSN\|SHISA5\|PRSS3\|A |

| | | | | | | | | |
|---|---|---|---|---|---|---|---|---|
| | | | | | | | | PS\|IGHA1\|ITIH4\|SERPINA3\|SERPINA1\|EGF\|FCGR3A\|HBB\|SPP1\|KNG1\|SEMG1\|SEMG2\|KLK1\|SOD3\|FLNA\|A1BG\|GNS\|AMBP\|DKFZP686I04196\|LAMP1\|ORM1\|RETN\|HPX\|CD59\|MGAM\|F2\|YIPF3\|PLA2G6\|IGKV3-20\|SH3BGRL3\|ORM2\|CDH11 |
| **6879** | 0.003727 | 0.042523 | 2 | 30 | 54 | 17786 | cellular iron ion homeostasis | TF\|HPX |
| **46824** | 0.003727 | 0.042523 | 2 | 30 | 54 | 17786 | positive regulation of nucleocytoplasmic transport | CDH1\|FLNA |
| **52547** | 0.0037764 | 0.042523 | 3 | 103 | 54 | 17786 | regulation of peptidase activity | TF\|F2\|CDH1 |
| **5770** | 0.0037764 | 0.042523 | 3 | 103 | 54 | 17786 | late endosome | LAMP1\|TF\|LAMP2 |
| **16485** | 0.0039859 | 0.04452 | 3 | 105 | 54 | 17786 | protein processing | TF\|MASP2\|PRSS3 |
| **1932** | 0.0042311 | 0.046881 | 4 | 217 | 54 | 17786 | regulation of protein amino acid phosphorylation | APOA1\|HPX\|F2\|EGF |
| **51049** | 0.0044709 | 0.04854 | 6 | 513 | 54 | 17786 | regulation of transport | APOA1\|F2\|CDH1\|EGF\|FLNA\|AHSG |
| **10740** | 0.0045863 | 0.04854 | 4 | 222 | 54 | 17786 | positive regulation of intracellular protein kinase cascade | TF\|HPX\|SHISA5\|FLNA |
| **7416** | 0.0047695 | 0.04854 | 2 | 34 | 54 | 17786 | synapse assembly | CDH1\|CDH2 |
| **30005** | 0.0048089 | 0.04854 | 4 | 225 | 54 | 17786 | cellular di-, tri-valent inorganic cation homeostasis | KNG1\|TF\|HPX\|F2 |
| **267** | 0.0048138 | 0.04854 | 9 | 1078 | 54 | 17786 | cell fraction | AMBP\|DKFZP686I04196\|LAMP1\|LAMP2\|CD59\|F2\|EGF\|IGHM\|SOD3 |
| **5886** | 0.0048913 | 0.04854 | 20 | 3731 | 54 | 17786 | plasma membrane | TF\|UMOD\|CDH1\|CDH2\|PIGR\|IGHM\|FLNA\|AMBP\|LAMP1\|AZGP1\|LAMP2\|APOA1\|CD59\|APS\|MGAM\|F2\|YIPF3\|EGF\|FCGR3A\|CDH11 |
| **51960** | 0.0049612 | 0.04854 | 4 | 227 | 54 | 17786 | regulation of nervous system development | F2\|CDH2\|EGF\|SPP1 |
| **31399** | 0.0049706 | 0.04854 | 5 | 366 | 54 | 17786 | regulation of protein modification process | APOA1\|HPX\|F2\|CDH1\|EGF |
| **5976** | 0.0050159 | 0.04854 | 3 | 114 | 54 | 17786 | polysaccharide metabolic process | GNS\|MGAM\|ITIH4 |
| **51604** | 0.0051393 | 0.04854 | 3 | 115 | 54 | 17786 | protein maturation | TF\|MASP2\|PRSS3 |
| **48518** | 0.0051395 | 0.04854 | 14 | 2207 | 54 | 17786 | positive regulation of biological process | KNG1\|TF\|MASP2\|CDH1\|FLNA\|AHSG\|APOA1\|HPX\|APS\|F2\|SHISA5\|EGF\|HBB\|SPP1 |
| **55072** | 0.0053354 | 0.04854 | 2 | 36 | 54 | 17786 | iron ion homeostasis | TF\|HPX |
| **5913** | 0.0053354 | 0.04854 | 2 | 36 | 54 | 17786 | cell-cell adherens junction | CDH1\|CDH2 |
| **4175** | 0.0053809 | 0.04854 | 5 | 373 | 54 | 17786 | endopeptidase activity | MASP2\|PRSS3\|F2\|APS\|KLK1 |
| **7155** | 0.005475 | 0.04854 | 7 | 711 | 54 | 17786 | cell adhesion | AMBP\|AZGP1\|LGALS3BP\|CDH1\|CDH2\|CDH11\|SPP1 |
| **22610** | 0.0055166 | 0.04854 | 7 | 712 | 54 | 17786 | biological adhesion | AMBP\|AZGP1\|LGALS3BP\|CDH1\|CDH2\|CDH11\|SPP1 |
| **51128** | 0.0056225 | 0.04854 | 6 | 538 | 54 | 17786 | regulation of cellular component organization | APOA1\|GSN\|CDH2\|EGF\|SPP1\|AHSG |
| **33013** | 0.0056293 | 0.04854 | 2 | 37 | 54 | 17786 | tetrapyrrole metabolic process | AMBP\|HPX |
| **6778** | 0.0056293 | 0.04854 | 2 | 37 | 54 | 17786 | porphyrin metabolic process | AMBP\|HPX |
| **35468** | 0.0058146 | 0.04854 | 5 | 380 | 54 | 17786 | positive regulation of signaling pathway | TF\|HPX\|SHISA5\|EGF\|FLNA |
| **4869** | 0.0059306 | 0.04854 | 2 | 38 | 54 | 17786 | cysteine-type endopeptidase inhibitor activity | KNG1\|AHSG |
| **5504** | 0.0059306 | 0.04854 | 2 | 38 | 54 | 17786 | fatty acid binding | AZGP1\|ALB |
| **55066** | 0.0060312 | 0.04854 | 4 | 240 | 54 | 17786 | di-, tri-valent inorganic cation homeostasis | KNG1\|TF\|HPX\|F2 |

| | | | | | | | | |
|---|---|---|---|---|---|---|---|---|
| **44004** | 0.0060631 | 0.04854 | 1 | 2 | 54 | 17786 | disruption by symbiont of host cells | ALB |
| **70508** | 0.0060631 | 0.04854 | 1 | 2 | 54 | 17786 | cholesterol import | APOA1 |
| **19836** | 0.0060631 | 0.04854 | 1 | 2 | 54 | 17786 | hemolysis by symbiont of host erythrocytes | ALB |
| **52331** | 0.0060631 | 0.04854 | 1 | 2 | 54 | 17786 | hemolysis of cells in other organism involved in symbiotic interaction | ALB |
| **52332** | 0.0060631 | 0.04854 | 1 | 2 | 54 | 17786 | modification by organism of cell membrane in other organism involved in symbiotic interaction | ALB |
| **19862** | 0.0060631 | 0.04854 | 1 | 2 | 54 | 17786 | IgA binding | AMBP |
| **8269** | 0.0060631 | 0.04854 | 1 | 2 | 54 | 17786 | JAK pathway signal transduction adaptor activity | APS |
| **10641** | 0.0060631 | 0.04854 | 1 | 2 | 54 | 17786 | positive regulation of platelet-derived growth factor receptor signaling pathway | TF |
| **35382** | 0.0060631 | 0.04854 | 1 | 2 | 54 | 17786 | sterol transmembrane transport | APOA1 |
| **35376** | 0.0060631 | 0.04854 | 1 | 2 | 54 | 17786 | sterol import | APOA1 |
| **50711** | 0.0060631 | 0.04854 | 1 | 2 | 54 | 17786 | negative regulation of interleukin-1 secretion | APOA1 |
| **48670** | 0.0060631 | 0.04854 | 1 | 2 | 54 | 17786 | regulation of collateral sprouting | SPP1 |
| **48671** | 0.0060631 | 0.04854 | 1 | 2 | 54 | 17786 | negative regulation of collateral sprouting | SPP1 |
| **51659** | 0.0060631 | 0.04854 | 1 | 2 | 54 | 17786 | maintenance of mitochondrion location | ALB |
| **71285** | 0.0060631 | 0.04854 | 1 | 2 | 54 | 17786 | cellular response to lithium ion | CDH1 |
| **52025** | 0.0060631 | 0.04854 | 1 | 2 | 54 | 17786 | modification by symbiont of host cell membrane | ALB |
| **52043** | 0.0060631 | 0.04854 | 1 | 2 | 54 | 17786 | modification by symbiont of host cellular component | ALB |
| **52111** | 0.0060631 | 0.04854 | 1 | 2 | 54 | 17786 | modification by symbiont of host structure | ALB |
| **52188** | 0.0060631 | 0.04854 | 1 | 2 | 54 | 17786 | modification of cellular component in other organism involved in symbiotic interaction | ALB |
| **52185** | 0.0060631 | 0.04854 | 1 | 2 | 54 | 17786 | modification of structure of other organism involved in symbiotic interaction | ALB |
| **44179** | 0.0060631 | 0.04854 | 1 | 2 | 54 | 17786 | hemolysis of cells in other organism | ALB |
| **1907** | 0.0060631 | 0.04854 | 1 | 2 | 54 | 17786 | killing by symbiont of host cells | ALB |
| **1897** | 0.0060631 | 0.04854 | 1 | 2 | 54 | 17786 | cytolysis by symbiont of host cells | ALB |
| **51715** | 0.0060631 | 0.04854 | 1 | 2 | 54 | 17786 | cytolysis of cells of another organism | ALB |
| **51801** | 0.0060631 | 0.04854 | 1 | 2 | 54 | 17786 | cytolysis of cells in other organism involved in symbiotic interaction | ALB |
| **50821** | 0.006239 | 0.049661 | 2 | 39 | 54 | 17786 | protein stabilization | APOA1\|FLNA |

corr p-value: Multiple testing correction using Benjamini&Hochberg (FDR) method, x: Number of genes in the given cluster annotated to a certain GO class, n: Number of genes in the reference set annotated to a certain GO class, X: Total number of genes in the given cluster N: Total number of genes in reference set.

**Supplemental Table S5: Over-representation of gene ontological terms observed in the down-regulated gene set identified in TB groups**

| GO-ID | p-value | corr p-value | x | n | X | N | Description | Genes in test set |
|---|---|---|---|---|---|---|---|---|
| **5576** | 1.32E-09 | 1.21E-06 | 16 | 2027 | 26 | 17790 | extracellular region | GC\|IGHG3\|RNASE2\|SPARCL1\|HSPG2\|B2M\|SERPINA7\|FGA\|PTGDS\|CLEC3B\|SERPINA5\|COL1A1\|NRG1\|CHGB\|WFDC2\|SPP1 |
| **5615** | 4.63E-08 | 1.54E-05 | 10 | 747 | 26 | 17790 | extracellular space | GC\|FGA\|SERPINA7\|CLEC3B\|HSPG2\|COL1A1\|NRG1\|WFDC2\|SPP1\|B2M |
| **44421** | 5.08E-08 | 1.54E-05 | 11 | 985 | 26 | 17790 | extracellular region part | GC\|FGA\|SERPINA7\|CLEC3B\|SPARCL1\|HSPG2\|COL1A1\|NRG1\|WFDC2\|SPP1\|B2M |
| **32501** | 1.27E-05 | 0.002521 | 17 | 4376 | 26 | 17790 | multicellular organismal process | GM2A\|AXL\|HSPG2\|CNTFR\|SDC4\|SERPINA7\|FGA\|CD44\|CLEC3B\|SERPINA5\|CSTB\|ROBO4\|KRT1\|COL1A1\|NRG1\|WFDC2\|SPP1 |
| **33273** | 1.38E-05 | 0.002521 | 4 | 102 | 26 | 17790 | response to vitamin | CD44\|SERPINA7\|COL1A1\|SPP1 |
| **50896** | 3.34E-05 | 0.004601 | 15 | 3633 | 26 | 17790 | response to stimulus | IGHG3\|RNASE2\|GM2A\|AXL\|B2M\|PTGDS\|SERPINA7\|FGA\|CD44\|CSTB\|KRT1\|COL1A1\|CREB3L3\|NRG1\|SPP1 |
| **9991** | 3.53E-05 | 0.004601 | 5 | 264 | 26 | 17790 | response to extracellular stimulus | CD44\|SERPINA7\|AXL\|COL1A1\|SPP1 |
| **4866** | 5.65E-05 | 0.005424 | 4 | 146 | 26 | 17790 | endopeptidase inhibitor activity | SERPINA7\|SERPINA5\|CSTB\|WFDC2 |
| **61135** | 5.81E-05 | 0.005424 | 4 | 147 | 26 | 17790 | endopeptidase regulator activity | SERPINA7\|SERPINA5\|CSTB\|WFDC2 |
| **30155** | 5.96E-05 | 0.005424 | 4 | 148 | 26 | 17790 | regulation of cell adhesion | COL1A1\|NRG1\|SDC4\|SPP1 |
| **10810** | 6.54E-05 | 0.005424 | 3 | 54 | 26 | 17790 | regulation of cell-substrate adhesion | COL1A1\|SDC4\|SPP1 |
| **30414** | 7.14E-05 | 0.005424 | 4 | 155 | 26 | 17790 | peptidase inhibitor activity | SERPINA7\|SERPINA5\|CSTB\|WFDC2 |
| **7584** | 9.75E-05 | 0.006842 | 4 | 168 | 26 | 17790 | response to nutrient | CD44\|SERPINA7\|COL1A1\|SPP1 |
| **9605** | 0.000117 | 0.007306 | 6 | 551 | 26 | 17790 | response to external stimulus | RNASE2\|CD44\|SERPINA7\|AXL\|COL1A1\|SPP1 |
| **61134** | 0.000122 | 0.007306 | 4 | 178 | 26 | 17790 | peptidase regulator activity | SERPINA7\|SERPINA5\|CSTB\|WFDC2 |
| **33189** | 0.00013 | 0.007306 | 3 | 68 | 26 | 17790 | response to vitamin A | CD44\|SERPINA7\|COL1A1 |
| **45785** | 0.000136 | 0.007306 | 3 | 69 | 26 | 17790 | positive regulation of cell adhesion | NRG1\|SDC4\|SPP1 |
| **42221** | 0.000146 | 0.0074 | 9 | 1465 | 26 | 17790 | response to chemical stimulus | RNASE2\|CD44\|FGA\|SERPINA7\|KRT1\|COL1A1\|CREB3L3\|SPP1\|B2M |
| **45177** | 0.000218 | 0.010453 | 4 | 207 | 26 | 17790 | apical part of cell | CD44\|GM2A\|NRG1\|SPP1 |
| **4867** | 0.00033 | 0.015028 | 3 | 93 | 26 | 17790 | serine-type endopeptidase inhibitor activity | SERPINA7\|SERPINA5\|WFDC2 |
| **31667** | 0.000401 | 0.017406 | 4 | 243 | 26 | 17790 | response to nutrient levels | CD44\|SERPINA7\|COL1A1\|SPP1 |
| **7275** | 0.000441 | 0.018298 | 12 | 2972 | 26 | 17790 | multicellular organismal development | CD44\|SERPINA7\|CLEC3B\|AXL\|HSPG2\|KRT1\|ROBO4\|CNTFR\|COL1A1\|NRG1\|SDC4\|SPP1 |
| **1568** | 0.000556 | 0.022038 | 4 | 265 | 26 | 17790 | blood vessel development | CD44\|HSPG2\|ROBO4\|COL1A1 |
| **1944** | 0.000621 | 0.023616 | 4 | 273 | 26 | 17790 | vasculature development | CD44\|HSPG2\|ROBO4\|COL1A1 |
| **4857** | 0.000674 | 0.024596 | 4 | 279 | 26 | 17790 | enzyme inhibitor activity | SERPINA7\|SERPINA5\|CSTB\|WFDC2 |
| **48856** | 0.000705 | 0.024724 | 11 | 2656 | 26 | 17790 | anatomical structure development | CD44\|CLEC3B\|AXL\|HSPG2\|KRT1\|ROBO4\|CNTFR\|COL1A1\|NRG1\|SDC4\|SPP1 |
| **2020** | 0.000758 | 0.025619 | 2 | 28 | 26 | 17790 | protease binding | SERPINA5\|CSTB |
| **48583** | 0.000857 | 0.027905 | 5 | 524 | 26 | 17790 | regulation of response to stimulus | PTGDS\|CD44\|KRT1\|SPP1\|B2M |
| **32502** | 0.000979 | 0.029173 | 12 | 3235 | 26 | 17790 | developmental process | CD44\|SERPINA7\|CLEC3B\|AXL\|HSPG2\|KRT1\|ROBO4\|CNTFR\|COL1A1\|NRG1\|SDC4\|SPP1 |

| | | | | | | | | |
|---|---|---|---|---|---|---|---|---|
| **9611** | 0.000989 | 0.029173 | 5 | 541 | 26 | 17790 | response to wounding | CD44\|FGA\|KRT1\|NRG1\|SPP1 |
| **10811** | 0.000992 | 0.029173 | 2 | 32 | 26 | 17790 | positive regulation of cell-substrate adhesion | SDC4\|SPP1 |
| **1649** | 0.001255 | 0.030997 | 2 | 36 | 26 | 17790 | osteoblast differentiation | COL1A1\|SPP1 |
| **31214** | 0.001255 | 0.030997 | 2 | 36 | 26 | 17790 | biomineral formation | COL1A1\|SPP1 |
| **30234** | 0.001256 | 0.030997 | 6 | 860 | 26 | 17790 | enzyme regulator activity | SERPINA7\|GM2A\|SERPINA5\|CSTB\|NRG1\|WFDC2 |
| **10033** | 0.001325 | 0.030997 | 6 | 869 | 26 | 17790 | response to organic substance | CD44\|SERPINA7\|COL1A1\|CREB3L3\|SPP1\|B2M |
| **48731** | 0.001421 | 0.030997 | 10 | 2422 | 26 | 17790 | system development | CD44\|CLEC3B\|AXL\|HSPG2\|KRT1\|ROBO4\|CNTFR\|COL1A1\|NRG1\|SPP1 |
| **2477** | 0.001462 | 0.030997 | 1 | 1 | 26 | 17790 | antigen processing and presentation of exogenous peptide antigen via MHC class Ib | B2M |
| **2481** | 0.001462 | 0.030997 | 1 | 1 | 26 | 17790 | antigen processing and presentation of exogenous protein antigen via MHC class Ib, TAP-dependent | B2M |
| **32428** | 0.001462 | 0.030997 | 1 | 1 | 26 | 17790 | beta-N-acetylgalactosaminidase activity | GM2A |
| **30290** | 0.001462 | 0.030997 | 1 | 1 | 26 | 17790 | sphingolipid activator protein activity | GM2A |
| **48685** | 0.001462 | 0.030997 | 1 | 1 | 26 | 17790 | negative regulation of collateral sprouting of intact axon in response to injury | SPP1 |
| **48683** | 0.001462 | 0.030997 | 1 | 1 | 26 | 17790 | regulation of collateral sprouting of intact axon in response to injury | SPP1 |
| **34238** | 0.001462 | 0.030997 | 1 | 1 | 26 | 17790 | macrophage fusion | CD44 |
| **30246** | 0.001805 | 0.037407 | 4 | 364 | 26 | 17790 | carbohydrate binding | CD44\|CLEC3B\|SERPINA5\|KRT1 |
| **48646** | 0.002012 | 0.040775 | 4 | 375 | 26 | 17790 | anatomical structure formation involved in morphogenesis | CD44\|HSPG2\|ROBO4\|COL1A1 |
| **48638** | 0.00241 | 0.047778 | 2 | 50 | 26 | 17790 | regulation of developmental growth | NRG1\|SPP1 |
| **33031** | 0.002921 | 0.048294 | 1 | 2 | 26 | 17790 | positive regulation of neutrophil apoptosis | CD44 |
| **33029** | 0.002921 | 0.048294 | 1 | 2 | 26 | 17790 | regulation of neutrophil apoptosis | CD44 |
| **2428** | 0.002921 | 0.048294 | 1 | 2 | 26 | 17790 | antigen processing and presentation of peptide antigen via MHC class Ib | B2M |
| **4667** | 0.002921 | 0.048294 | 1 | 2 | 26 | 17790 | prostaglandin-D synthase activity | PTGDS |
| **48670** | 0.002921 | 0.048294 | 1 | 2 | 26 | 17790 | regulation of collateral sprouting | SPP1 |
| **48671** | 0.002921 | 0.048294 | 1 | 2 | 26 | 17790 | negative regulation of collateral sprouting | SPP1 |

| | | | | | | | | |
|---|---|---|---|---|---|---|---|---|
| **5584** | 0.002921 | 0.048294 | 1 | 2 | 26 | 17790 | collagen type I | COL1A1 |
| **60346** | 0.002921 | 0.048294 | 1 | 2 | 26 | 17790 | bone trabecula formation | COL1A1 |
| **5889** | 0.002921 | 0.048294 | 1 | 2 | 26 | 17790 | hydrogen:potassium-exchanging ATPase complex | GM2A |
| **42060** | 0.002965 | 0.048294 | 3 | 199 | 26 | 17790 | wound healing | CD44\|FGA\|NRG1 |
| **48513** | 0.003098 | 0.049567 | 8 | 1792 | 26 | 17790 | organ development | CD44\|AXL\|HSPG2\|KRT1\|ROBO4\|COL1A1\|NRG1\|SPP1 |

**Note**
corr p-value: Multiple testing correction using Benjamini&Hochberg (FDR) method
x: Number of genes in the given cluster annotated to a certain GO class
n: Number of genes in the reference set annotated to a certain GO class
X: Total number of genes in the given cluster
N: Total number of genes in reference set
